# Analysis of 6.4 million SARS-CoV-2 genomes identifies mutations associated with fitness

**DOI:** 10.1101/2021.09.07.21263228

**Authors:** Fritz Obermeyer, Martin Jankowiak, Nikolaos Barkas, Stephen F. Schaffner, Jesse D. Pyle, Lonya Yurkovetskiy, Matteo Bosso, Daniel J. Park, Mehrtash Babadi, Bronwyn L. MacInnis, Jeremy Luban, Pardis C. Sabeti, Jacob E. Lemieux

## Abstract

Repeated emergence of SARS-CoV-2 variants with increased fitness necessitates rapid detection and characterization of new lineages. To address this need, we developed PyR_0_, a hierarchical Bayesian multinomial logistic regression model that infers relative prevalence of all viral lineages across geographic regions, detects lineages increasing in prevalence, and identifies mutations relevant to fitness. Applying PyR_0_ to all publicly available SARS-CoV-2 genomes, we identify numerous substitutions that increase fitness, including previously identified spike mutations and many non-spike mutations within the nucleocapsid and nonstructural proteins. PyR_0_ forecasts growth of new lineages from their mutational profile, identifies viral lineages of concern as they emerge, and prioritizes mutations of biological and public health concern for functional characterization.

**One Sentence summary:** A Bayesian hierarchical model of all SARS-CoV-2 viral genomes predicts lineage fitness and identifies associated mutations.

## Main Text

The SARS-CoV-2 pandemic has been characterized by repeated waves of cases driven by the emergence of new lineages with higher fitness, where fitness encompasses any trait that affects the lineage’s growth, including its basic reproduction number (R_0_), ability to evade existing immunity, and generation time. Rapidly identifying such lineages as they emerge and accurately forecasting their dynamics is critical for guiding outbreak response. Doing so effectively would benefit from the ability to interrogate the entirety of the global SARS-CoV-2 genomic dataset. The large size (currently over 7.5 million virus genomes) and geographic and temporal variability of the available data present significant challenges that will only become greater as more viruses are sequenced. Current phylogenetic approaches are computationally inefficient on datasets with more than ∼5000 samples and take days to run at that scale. Ad hoc methods to estimate the relative fitness of particular SARS-CoV-2 lineages are a computationally efficient alternative (*1–3*), but have typically relied on models in which one or two lineages of interest are compared to all others and do not capture the complex dynamics of multiple co-circulating lineages.

Furthermore, estimates of relative fitness based on lineage frequency data alone (*2–4*) do not take advantage of additional statistical power that can be gained from analyzing the independent appearance and growth of the same mutation in multiple lineages. Performing a mutation-based analysis of lineage prevalence has the additional advantage of identifying specific genetic determinants of a lineage’s phenotype, which is critically important both for understanding the biology of transmission and pathogenesis and for predicting the phenotype of new lineages. The SARS-CoV-2 pandemic has already been dominated by several genetic changes of functional and epidemiological importance, including the spike (S) D614G mutation that is associated with higher SARS-CoV-2 loads (*5, 6*). In addition, mutations found in Variants of Concern (VoC), such as S:N439R, S:N501Y, and S:E484K, have been linked, respectively, to increased transmissibility (*7*), enhanced binding to ACE2 (*8*), and antibody escape (*9, 10*). Despite these successes, identifying functionally important mutations in the context of a large background of genetic variants of little or no phenotypic consequence remains challenging.

We set out to formulate a principled approach to modeling the relative fitness of SARS-CoV-2 lineages, estimating their growth as a linear combination of the effects of individual mutations. We developed PyR_0_, a hierarchical Bayesian regression model that enables scalable analysis of the complete set of publicly available SARS-CoV-2 genomes, and that could be applied to any viral genomic dataset and to other phenotypes. The model, which is summarized in Figure 1A and described in detail in the supplemental note, avoids the complexity of full phylogenetic inference by first clustering genomes by genetic similarity (refining PANGO lineages (*11*)), and then estimating the incremental effect on growth rate of each of the most common amino acid changes on the lineages in which they appear. By regressing growth rate as a function of genome sequence, the model shares statistical strength among genetically similar lineages without explicitly relying on phylogeny. By modeling only the multinomial proportion of different lineages rather than the absolute number of samples for each lineage (*4, 12*), and by doing so within 14-day intervals in 1560 globally distributed geographic regions, the model achieves robustness to a number of sources of bias that affect all lineages, across regions, and over time, including differences in data collection and changes in transmission due to such factors as social behavior, public health policy, and vaccination.

**Figure 1.**
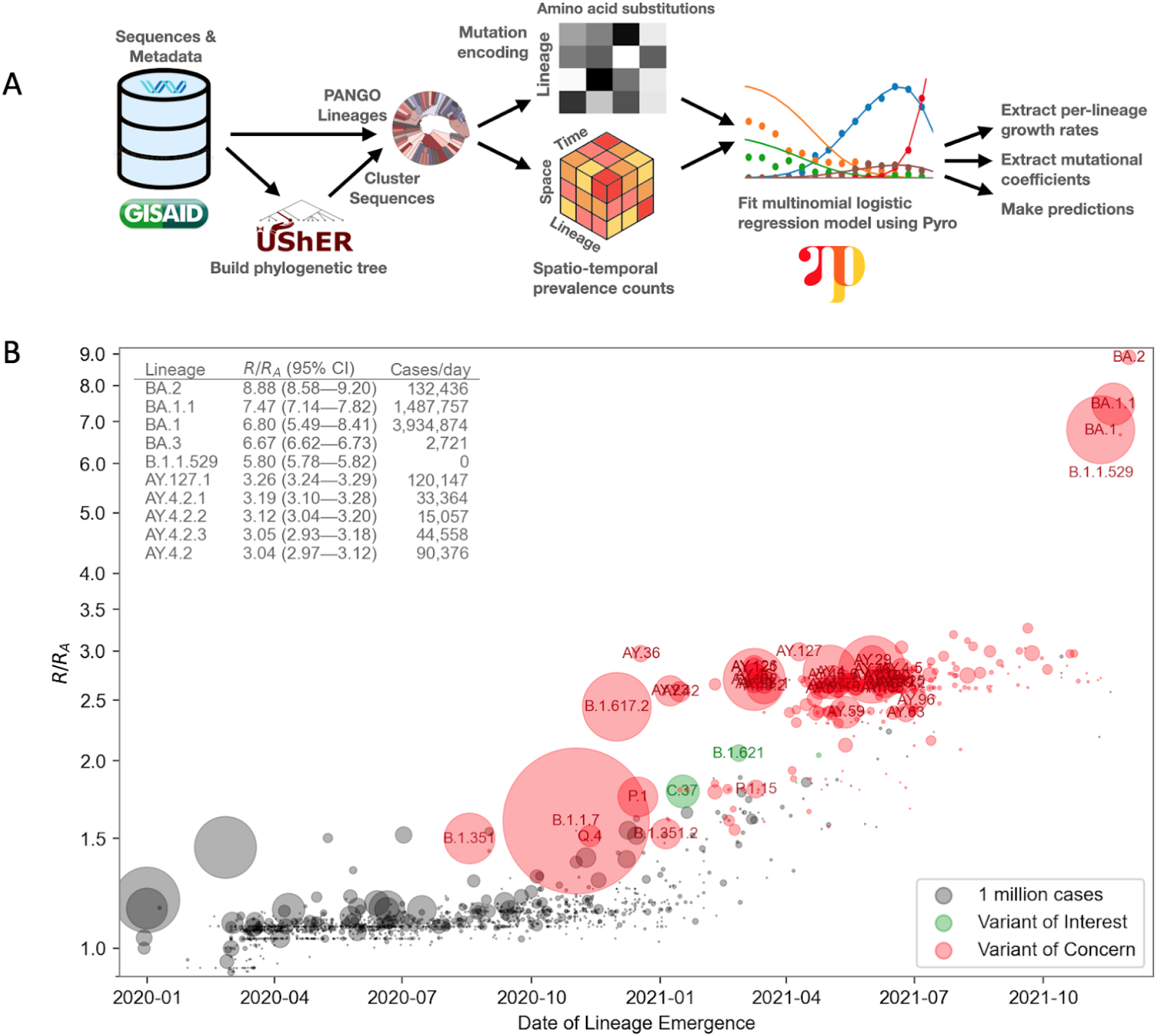
**A.** Overview of the PyR_0_ analysis pipeline. After clustering UShER’s mutation annotated tree, sequence data are used to construct spatio-temporal lineage prevalence counts y_tpc_ and amino acid substitution covariates X_cf_. Pyro is used to fit a Bayesian multivariate logistic multinomial regression model to y_tpc_ and X_cf_. **B.** Relative fitness versus date of lineage emergence. Circle size is proportional to cumulative case count inferred from lineage proportion estimates and confirmed case counts. Inset table lists the 10 fittest lineages inferred by the model. R/R_A_ is the fold increase in relative fitness over the Wuhan (A) lineage, assuming a fixed generation time of 5.5 days.

We fit PyR_0_ to 6,466,300 SARS-CoV-2 genomes available on GISAID (*13, 14*) as of January 20, 2022, in a model that contained 1544 PANGO lineages and 2904 nonsynonymous mutations. The output of the model is a posterior distribution for the relative fitness (exponential growth rate) of each lineage and for the contribution to the fitness from each mutation. Fitting this large model is computationally challenging, so we used stochastic variational inference, an approximate inference method that reduced our task to solving a 75-million-dimensional optimization problem on a GPU. Inference was implemented in the Pyro (*15*) probabilistic programming framework (see Supplemental Materials). The trained model can be used to infer lineage fitness, predict the fitness of completely new lineages, forecast future lineage proportions, and estimate the effects of individual mutations on fitness.

The model’s lineage fitness estimates (Figure 1B) show a modest upward trend over time among all lineages, accompanied by numerous lineages with dramatically higher fitness. Sensitivity analyses revealed broad consistency of fitness estimates across spatial data subsets (Figure S1). The upward trend may in part reflect an upward bias caused by the lineage assignment process, as can be seen in simulation studies (Figure S2), but the high tail of the distribution exhibits elevated fitness values far in excess of this trend. The rate of increase in fitness was not constant between the emergence of the virus into human populations in late 2019 and early 2022. Rather, periods of rapid evolution in fitness occurred and heralded new waves of increase in case counts (Figure 1B and Figure 2CDE). The model correctly inferred BA.2 to have the highest fitness to date, 8.9-fold (95% CI, 8.6-9.2) higher than the original A lineage (Figure 1B inset). Similar fitness was estimated for other Omicron sub-lineages BA.1 and BA.1.1 (Figure 1B). These fitness estimates, obtained in mid January 2022, predict B.1.1.529 and sublineages (collectively called Omicron in the WHO classification) will continue to displace other lineages, including the previously dominant Delta (Figure S3). While PANGO lineages facilitate communication by providing a stable nomenclature, we observed some PANGO lineages with multiple successive peaks in some regions, which could not be accounted for by a multivariate logistic growth model. We therefore algorithmically refined the 1544 PANGO lineages into 3000 finer clusters, and found our model identified significant heterogeneity within some PANGO lineages (Figure S4). Notably, B.1.1 displayed the greatest variability among lineages, followed by B.1.

**Figure 2.**
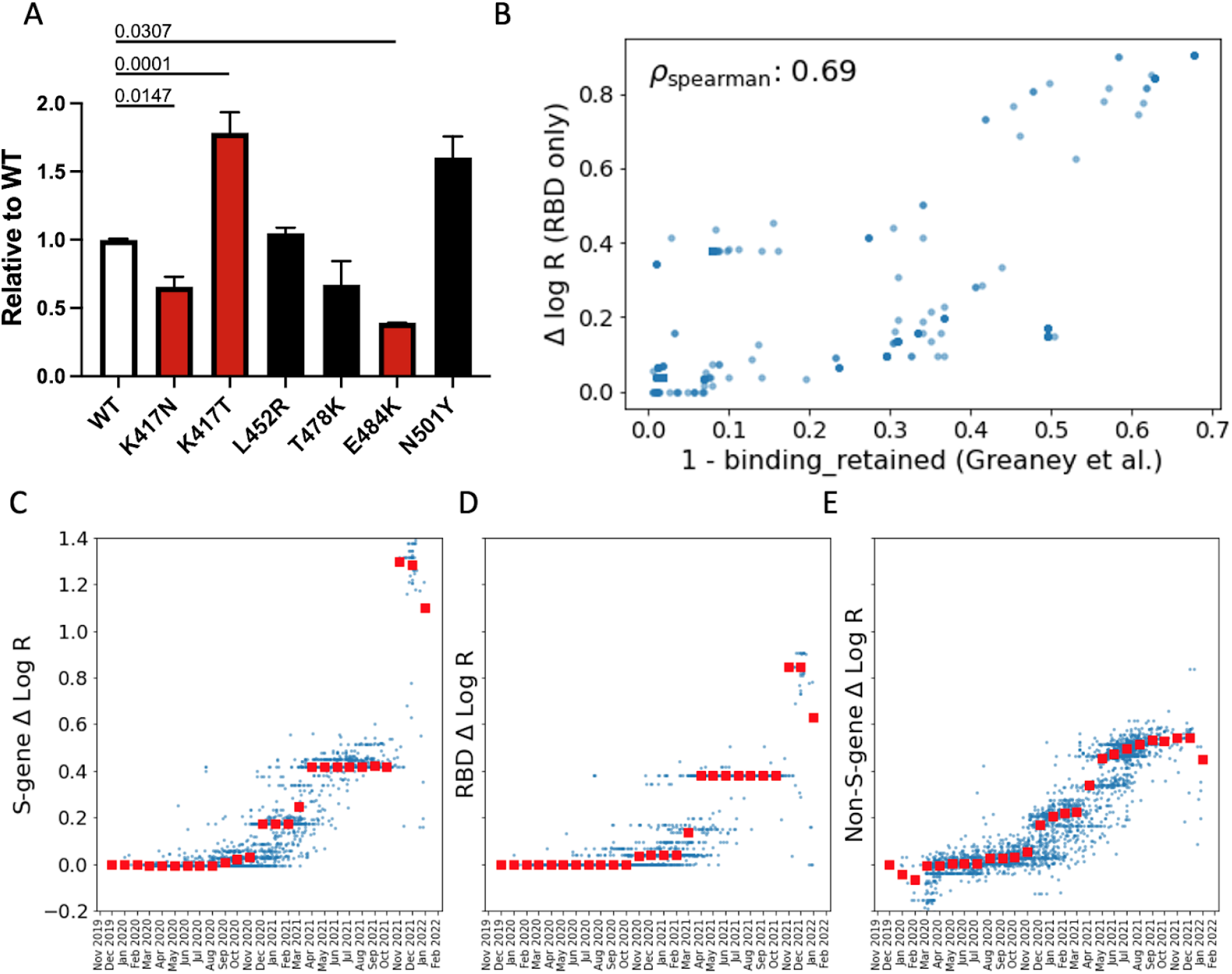
**A.** Infectivity relative to WT of lentiviral vectors pseudotyped with the indicated Spike mutants. Target cells were HEK293T cells expressing ACE2 and TMPRSS2 transgenes. The genetic background of the Spike was Wuhan-Hu-1 bearing D614G. Red bars were significantly different from WT (adjusted p values shown). Black bars were not significantly different from WT. **B.** For the 1701 SARS-CoV-2 clusters with at least one amino acid substitution in the RBD domain we compare: i) the PyR_0_ prediction for the contribution to Δ log R from RBD substitutions only; to ii) antibody binding computed using the antibody-escape calculator in (17). The escape calculator is based on an intuitive non-linear model parameterized using deep mutational scanning data for 33 neutralizing antibodies elicited by SARS-CoV-2. PyR_0_ predictions exhibit high (Spearman) correlation with predictions from Greaney et al. **C-E**. We dissect PyR_0_ Δ log R estimates into S-gene (C), RBD (D), and non-S-gene (E) contributions for 3000 SARS-CoV-2 clusters (blue dots). The horizontal axis corresponds to the date at which each cluster first emerged. Red squares denote the median Δ log R within each monthly bin. The increased importance of S-gene mutations (notably in the RBD) over non-S-gene mutations starting around November 2021 is apparent.

We found that the model would have provided early warning of the rise of VoCs had it been routinely applied to SARS-CoV-2 samples, highlighting the benefit of timely publication of genomic data. For example, PyR_0_ would have forecast the coming dominance of B.1.1.7 in late November 2020 (Figure S5A), while the first models forecasting its rapid rise were published in mid December 2020 (*16*). Similar predictions would have been available for BA.1 by early December 2021 (Figure S5B, S6) and for AY.4 by May 2021 (Figure S5C). Likewise the elevated fitness of BA.2 was identified by mid December 2021 on the basis of 76 observed sequences (Figure S6). While variant-specific models were accurate and useful (*2*) in predicting the rise of these lineages, each modeling effort was specific to a particular lineage and geographic region; by contrast, PyR_0_’s global approach provides similar early detection while also offering automated, rapid, and unbiased consideration of all variants and lineages, together with ranking based on relative fitness. When we tested the model’s predictive ability (Figure S5), we found that forecasts were reliable for 1-2 months into the future, when they tended to be disrupted by the emergence of a completely new strain (Table S1, Figure S7). Remarkably, the accuracy of forecasts stabilized typically within two weeks after the emergence of a new competitive lineage in a region (Figure S7).

By basing fitness estimates on the contributions of individual mutations, PyR_0_ can forecast the fitness of novel or hypothetical lineages using their mutational profiles alone. This is possible with SARS-CoV-2 because of the high rate of convergent evolution (Table 1, Figure S8), which allows the model to infer the fitness of new constellations of mutations based on the trajectories of other lineages in which they have previously emerged. This predictive capability is highly desirable from a public health standpoint because forecasts are available as soon as sequences from new lineages appear. To test the reliability of this kind of estimate, we fit leave-one-out estimators on subsets of the dataset with entire PANGO lineages removed (Figure S9). These estimators showed excellent agreement with estimators based on the observed behavior of the lineages, and they were also more accurate than naive phylogenetic estimators that assume the fitness of each new strain is equal to its parent lineage’s fitness (Pearson’s ρ = 0.983, after correcting for parent fitness, Figure S9). These results demonstrate the feasibility of this kind of estimate using the simplest possible linear-additive model, and provide a foundation for future research for more complex modeling that includes effects such as epistasis between mutations and migration across regions.

**Table 1:** Amino acid substitutions most significantly associated with increased fitness. Significance is defined as posterior mean / posterior standard deviation. Fitness is per 5.5 days (estimated generation time of the Wuhan (A) lineage (1, 19)). Final column: number of PANGO lineages in which each substitution emerged independently.

| Rank | Gene | Substitution | Fold Increase in Fitness | Number of Lineages |
| --- | --- | --- | --- | --- |
| 1 | S | H655Y | 1.051 | 33 |
| 2 | S | T95I | 1.046 | 30 |
| 3 | ORF1a | P3395H | 1.039 | 5 |
| 4 | S | N764K | 1.040 | 6 |
| 5 | ORF1a | K856R | 1.039 | 2 |
| 6 | S | S371L | 1.041 | 3 |
| 7 | E | T9I | 1.040 | 5 |
| 8 | S | Q954H | 1.040 | 5 |
| 9 | ORF9b | P10S | 1.039 | 25 |
| 10 | S | L981F | 1.040 | 2 |
| 11 | N | P13L | 1.040 | 25 |
| 12 | S | G339D | 1.039 | 4 |
| 13 | S | S375F | 1.040 | 5 |
| 14 | S | S477N | 1.039 | 47 |
| 15 | S | N679K | 1.040 | 11 |
| 16 | S | S373P | 1.040 | 5 |
| 17 | M | Q19E | 1.039 | 5 |
| 18 | S | D796Y | 1.038 | 11 |
| 19 | S | N969K | 1.040 | 5 |
| 20 | S | T547K | 1.038 | 3 |

Unbiased, genome-wide estimates of the effect of SARS-CoV-2 mutations on fitness also provide a powerful tool for better understanding the biology of fitness. Our model allowed us to estimate the contribution of 2904 amino acid substitutions (Figure 3a, Table 1) to lineage fitness and to rank them by inferred statistical significance (Figure S10). Cross-validation confirmed that these results replicate across different geographic regions (Figure S11). The highest concentrations of fitness-associated mutations were found in the S, N, and the ORF1 polyprotein genes (ORF1a and ORF1b, Figures 3A-B, S12-S13). Using spatial autocorrelation as a measure of spatial structure, we found evidence of functional hotspots in the S, N, ORF7a, ORF3a, and ORF1a genes (Table S2). Within S, there were three hotspots of fitness-enhancing mutations, each within a defined functional region: the N-terminal domain, the receptor-binding domain (RBD), and the furin-cleavage site (Figure 3B). We assessed mutational enrichment in the top-ranked set of mutations and identified an enrichment for lysine to asparagine mutations in the S gene (Figure S14C). We visualized top scoring mutations within atomic structures for the spike protein (Figure 3D-E), the nucleocapsid’s N-terminal domain (Figure 3F), the polymerase (Figure S15), and two proteases (Figure S16). Many of the top mutations in the S gene occurred in the receptor binding domain (RBD) making direct contacts with the ACE2 receptor, including K417N/T and E484K (Figures 3D-E). Two top-ranked mutations, T478K and S477N, occur in a flexible loop adjacent to the S-ACE2 interface (Figure 3E), suggesting that these mutations may affect the kinetics of receptor engagement and possibly viral entry. Other mutations occurred in regions proximal to essential enzymatic active sites of the viral replication (Figure S15) or protein processing (Figure S16) machinery.

**Figure 3.**
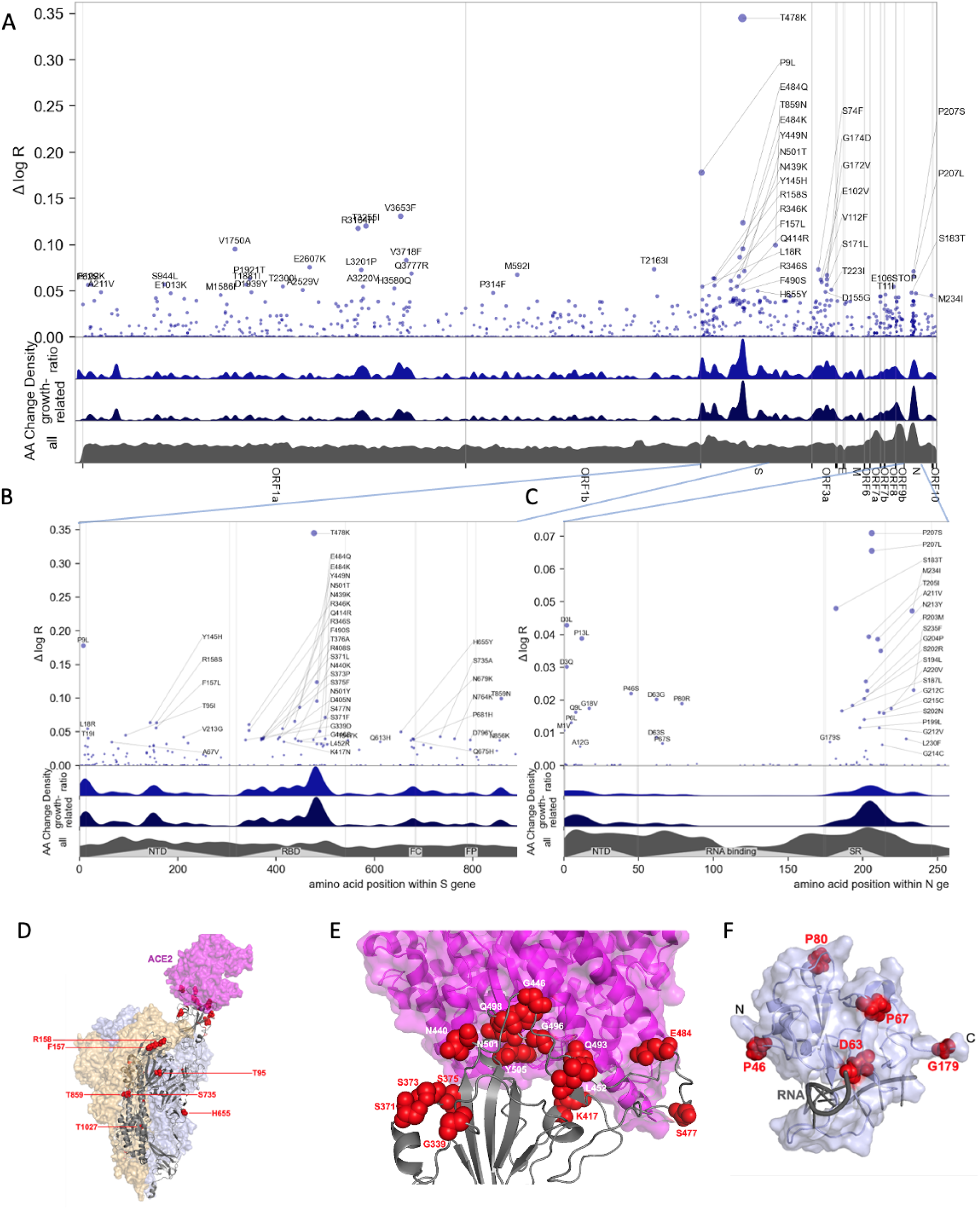
Manhattan plot of amino acid changes assessed in this study. **A.** Changes across the entire genome. **B.** Changes in the first 850 amino acids of S. In each of A-C the y axis shows effect size Δ log R, the estimated change in log relative fitness due to each amino acid change. The bottom three axes show the background density of all observed amino acid changes, the density of those associated with growth (weighted by |Δ log R|), and the ratio of the two. The top 55 amino acid changes are labeled. See Figure S13 for detailed views of S, N, ORF1a, and ORF1b. **C.** Changes in the first 250 amino acids of N. **D.** Structure of the spike-ACE2 complex (PDB: 7KNB). Spike subunits colored light blue, light orange, and gray. Top-ranked mutations are shown as red spheres. ACE2 is shown in magenta. **E.** Close-up view of the RBD interface. **F.** Top-ranked mutations in the N-terminal RNA-binding domain of N. Residues 44-180 of N (PDB: 7ACT) are shown in light blue. Amino acid positions corresponding to top mutations in this region are shown as red spheres. A 10-nt bound RNA is shown in gray.

We tested several of the high-scoring mutations in single-cycle infectivity assays as done previously (*6*), focusing on the RBD (Figure 2A). We found that while some individual mutations increased infectivity, on average high-scoring RBD mutations did not promote infectivity per se. We considered an alternate possibility that fitness of Spike mutations is driven by immune escape. Using RBD-aggregated mutations as a proxy for immune escape, we found that the fitness effect of these Spike mutations correlates well with antibody escape estimates from Greaney et al.(*17*) (Figure 2B). Together with the observed jump in fitness beginning in late 2021 (Figure 2C) associated with Spike mutations, but not mutations elsewhere in the genome (Figure 2E), these results suggest that immune escape is currently the dominant driver of fitness increases. In contrast to mutations in Spike, those in the serine-arginine rich region of N were linked to increased efficiency of SARS-CoV-2 genomic RNA packaging (*18*). Within ORF1, we found fitness-associated mutations across all viral enzymes, and clusters within additional non-structural proteins (nsps). The highest concentration of fitness-associated mutations is found in nsp4, nsp6, and nsp12–14 (Figure S12B,S13C-D), suggesting unexplored function at those sites. For example, nsp4 and nsp6 have roles in assembly of replication compartments, and substitutions in these regions may influence the kinetics of replication (see Supplemental Note 3). We note that while convergent evolution makes it possible to identify candidate functional mutations, observational data alone is insufficient to declare mutations as causal rather than merely correlated. For this reason hits identified by our study require functional followup, and can be prioritized by our uncertainty-ranked list of important mutations.

In summary, PyR_0_ provides an unbiased, automated approach for detecting viral lineages with increased fitness. By combining a model-based assessment of lineage fitness with absolute case counts, our model provides a global picture of the events of the first two years of the pandemic. Because it assesses the contribution of individual mutations and aggregates across all lineages and geographic regions, it can identify mutations and gene regions that likely increase fitness, and it can predict the relative fitness of new lineages based solely on viral sequence. Applied to the full set of publicly available SARS-CoV-2 genomes, it provides a principled, unbiased analysis of the mutations driving increased fitness of the virus, identifying experimentally established driver mutations in S and highlighting the key role of non-S mutations, particularly in N, ORF1b, and ORF1a, which have received relatively less research attention. By jointly estimating lineage and mutational fitness from millions of viral sequences across thousands of regions, PyR_0_ shares statistical strength across regions and mutations to yield mechanistic insight into viral fitness and enhance public health by forecasting lineage dynamics.

## Data Availability

All data was gathered from other public resources. Data preprocessing scripts are open source.

https://gisaid.org

https://github.com/CSSEGISandData/COVID-19

https://cov-lineages.org/

## Supplementary Materials

### Materials and methods

#### Data and Code Availability

Source code for data preprocessing and modeling and available at https://github.com/broadinstitute/pyro-cov. GISAID sequence data is publicly available at https://gisaid.org. PANGO lineage aliases are available at https://cov-lineages.org with source code at https://github.com/cov-lineages/lineages-website and lineage aliases available at https://github.com/cov-lineages/pango-designation. UShER phylogenies of public data are available at http://hgdownload.soe.ucsc.edu/goldenPath/wuhCor1/UShER_SARS-CoV-2. The whole genome map is available as part of NextClade at https://github.com/nextstrain/ncov/blob/50ceffa/defaults/annotation.gff. Structures of ORFab regions are available at https://www.ncbi.nlm.nih.gov/protein.

#### Regression model of relative fitness (PyR_0_ model)

We fit a Bayesian, hierarchical multinomial logistic regression model to data from GISAID using Pyro. Details are provided in the supplemental note below.

#### Simulation of lineages

We carried out a simulation study to determine whether the process of clustering genomes into named lineages could generate an artifactual increase in estimated fitness. The simulation was of a single neutrally evolving viral population with discrete generations and a stochastic population size generated by a highly overdispersed negative binomial distribution with the current fitness. (Overdispersion parameter = 0.11, which yields 10% of cases causing 80% of transmission.) The fitness is 2.5 for the first 10 generations; subsequently it drops to 1.5 until the viral population reaches 80,000 infections, whereupon it drops again to 0.8. When the population decreases to 10,000, the growth switches back to 1.5, and continues cycling when the high and low population thresholds are reached. (A model with a roughly constant-sized population yields similar results.) The population starts as a single named lineage. Each generation, the most successful nodes in that generation are determined by looking ahead four generations and counting descendants. New lineages are assigned to the nodes with the most descendants (minimum of 200 descendants), up to a maximum of 10 lineages per generation. 10% of all infections are randomly sampled and any lineage with fewer than 20 descendants is discarded. When all new lineages have been generated and all nodes assigned a lineage, a global multinomial logistic regression is performed, using the Python package sklearn.linear_model, yielding relative fitness estimates of all lineages.

#### Spatial analysis of mutation coefficients

To assess the spatial structure of the inferred amino acid coefficients β_f_ (described in Probabilistic Model below), we utilize the Moran I spatial autocorrelation statistic. We report (see Table S1) one-sided p-values for Moran I computed using a permutation test with 999,999 random permutations. We use a gaussian weighting function of the form exp(-distance^2^/lengthscale^2^), where distance is measured in units of nucleotides. We compute Moran I statistics both for individual genes and the entire genome. For larger genes whose extent is 1000+ nucleotides we use a length scale of 50 nucleotides. For smaller genes (e.g. ORF8) we set the length scale to one twentieth of the extent of the gene. When considering the entire set of amino acid changes, i.e. all 2,904 coefficients that make up β_f_, we compute the Moran I statistic for two different length scales. We note, however, that the Moran I statistic is somewhat simplistic, since it is designed to pick up spatial structure at a single length scale. In particular it can be insensitive to complex spatial structure that involves multiple overlapping substructures at different scales. Nevertheless it offers a simple quantitative metric for identifying spatial structure in the coefficients β_f_.

#### Analysis of substitution statistics

To assess enrichment of amino acid changes we compared the event frequencies for the leading mutation sets (as determined by posterior mean/std ranking) against a background of all mutations used as features in the model using multiple testing corrected binomial tests. We performed this analysis for both the asymmetric case (where A->V and V->A are different events) and for the symmetric case.

#### Comparison to other regression models

We fit logistic regression models in R version 4.0.3. The stats::glm() was used to fit binomial logistic regression models and the nnet::multinom function was used to fit multinomial logistic regression models. For multinomial logistic regressions, the data were filtered to contain sequences between January 1 2021 and December 31 2021 from the most common 25 pango lineages in the 10 countries with the most sequences available. The resulting dataset was downsampled to 10% of its initial size.

### Supplemental Note 1: Detailed description of PyR_0_ model

#### Data Preparation

We downloaded 6,466,300 samples from GISAID (*13, 14*) on January 20, 2021. Each sample record includes labels for time, location, PANGO lineage annotation (*11*), and genetic sequence. We discard records with missing time, location, or lineage. We use UShER (*20*) to build a mutation-annotated phylogenetic tree, discarding sequences whose alignment quality is not reported as “good”. We bin time intervals into 14-day segments, choosing a multiple of 7 to minimize weekly seasonality, but binning coarser than a week so as to reduce memory requirements; this results in 56 time bins.

Because sample counts vary widely across GISAID geographic region (by as much as five orders of magnitude), we aggregate regions into the following coarse partitions: each country counts as a region, and any first level subregion of a country counts as a region if it has at least 50 samples; otherwise it is aggregated into a whole-country bin. Note this means that e.g. a country may be split up into its larger regions, with smaller regions being subsumed into an aggregate country level bin. We then drop regions without samples in at least two different time intervals, resulting in 1560 regions in total. Figure S17 shows the distribution of samples among countries and GISAID regions.

After preprocessing, the model input data are a T × P × C = 56 × 1560 × 3000 shaped array y_tpc_ ∈ ℕ of counts (this array is sparse and our inference code uses a sparse representation), and an C × F = 3000 × 2904 shaped array X_sf_ ∈ {0, 1} of mutation features.

Cases per day (see Figure 3 inset) were estimated by multiplying confirmed case count data from Johns Hopkins University by the estimated proportion of each lineage within each (time, region) bin. We manually matched each GISAID region to the finest enclosing JHU region.

#### Lineage Clustering

Our method relies on a partitioning of genetic samples into clusters, where we estimate the fitness of each cluster. We initially tried to use the 1544 PANGO lineages as clusters, but found that some PANGO lineages appeared to include multiple distinct viruses of different fitness, e.g. B.1.1. exhibits two peaks in relative abundance in England, contrary to our multivariate logistic growth model. We therefore refined the 1544 PANGO lineages into 3000 finer clusters, with rates estimated individually for each cluster. Indeed Figure S4 shows that some PANGO lineages contain multiple distinct clusters of fitness estimates differing by more than a factor of two.

To create genetic clusters finer than PANGO lineages we began with a complete 4,833,238 node phylogeny of all GISAID samples maintained by Angie Hinrichs (*20*) (this phylogeny was created using UShER (*21*), excluding private mutations, masking difficult-to-sequence regions, eliding deletions, parsimoniously imputing missing sequence data). To coarsen the 4,833,238-node phylogenetic tree down to 3000 nodes (treated as clusters) we greedily collapsed parent-child edges, minimizing the the following distance function *TreeDistance*(-,-) between two mutation annotated trees

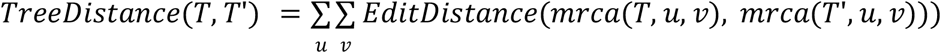

where T is the true mutation annotated tree, T’ is the collapsed tree whose nodes we treat as clusters, u and v are sample sequences, *mrca*(T,u,v) is the sequence of the most recent common ancestor of u,v in the mutation annotated tree T, and *EditDistance*(-,-) counts amino acid substitutions between two sequences. This objective function minimizes the mean edit distance between the true mrca sequence and its cluster’s sequence, for each pair of sequences. Changes in the objective function can be computed cheaply, and the O(n log(n)) time greedy algorithm can process the entire n=4,833,238 node phylogeny in under 5 minutes. Empirically this heuristic clustering produces trees that are approximately balanced in both cluster size and cluster-cluster edit distance, on both the true data and on synthetic datasets. Figure S18 shows the distribution of samples among both coarse PANGO lineages and the finer clusters. Figure S19 shows small example trees produced by clustering large synthetic trees.

#### Probabilistic Model

We model relative lineage growth with a hierarchical Bayesian regression model with a multinomial likelihood. Arrays in the model index over one or more indices: T=56 time steps (increments of 14 days) t; C=3000 clusters c; P=1560 regions (“places”) p; and F=2904 amino acid substitutions (“features”) f. The model, shown below, regresses lineage counts y_tpc_∈ ℕ in each time-region-lineage bin against amino acid mutation covariates X_cf_ ∈ {0,1}. The variables y and X are observed and all other variables in the model are latent. Each latent variable is governed by a prior distribution. The full model is specified as follows (visualized in Figure S20), where the observed counts y_tpc_ are underlined:

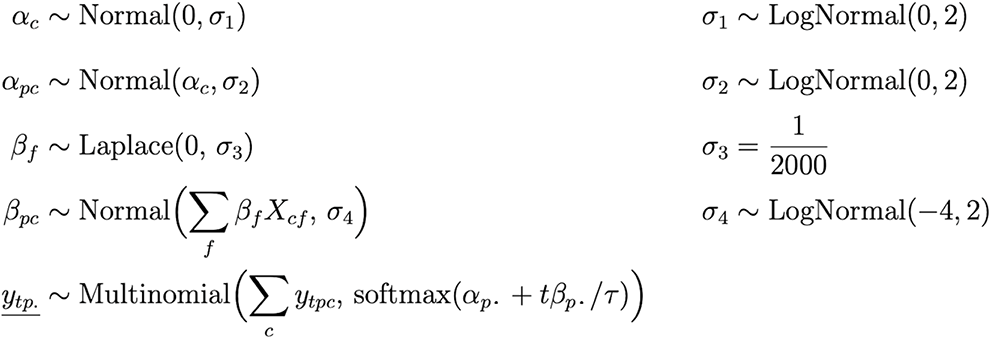

The proportion of lineages in a single time-region bin is modeled as a Multinomial distribution whose probability parameter is a multivariate logistic growth function softmax(α_p**·**_ + tβ_p**·**_/τ) with intercept α_pc_ and slope β_pc_ in units of generation time τ = 5.5 days (these units are for interpretability only; the model does not use the notion of generation, and thus is robust to changes in generation time). Here the dot subscripts α_p**·**_∈ ℝ^C^, β_p**·**_∈ ℝ^C^, and y_tp._∈ ℕ^C^ denote vectors over cluster ids. The softmax function implements the multivariate generalization of logistic growth, inputting and outputting vectors, and is defined as

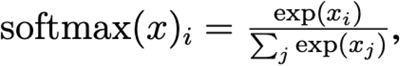

For a simple model of two lineages, each of the two components of the softmax function are sigmoid curves; however for more lineages, the functional forms may be more complex. Early iterations of the model used overdispersed likelihoods such as Dirichlet-Multinomial to account for additional variability not directly encoded in the generative process. However, we found that we can obtain much more accurate model predictions by using a Multinomial likelihood and accounting for model misfit by adding hierarchical structure elsewhere. The intercepts α_pc_ denote initial relative log prevalence of cluster c in region p; these are modeled hierarchically around the global relative log prevalence α_c_ of each cluster. The slopes β_pc_ are modeled hierarchically around global per-cluster fitness Σ*_f_* β*_f_ X*c*_f_* that are linearly regressed against amino acid substitution features X_cf_. These linear coefficients β_f_ can be directly interpreted as the effect of a mutation on a lineage’s fitness, all other variation being equal. In figures we plot posterior means 𝔼[β*_f_* |*data*] =: Δ log *R* as an estimate of effect size and plot the posterior z-score |𝔼[β*_f_* |*data*]| / 𝕍 [β*_f_* |*data*]^1/2^ =: |µ|/σ as a proxy for statistical significance.

Note that by regressing against amino acid changes we obviate the need to directly incorporate phylogenetic information into the model: if two lineages are close together in a phylogeny, then their amino acid features are likely also similar, so their regressed fitness values will likely be similar. By sharing statistical strength in this way we are also able to make accurate predictions for emergent lineages with few observations. (Note phylogenetic information is still used in preprocessing, since our clustering is created from an UShER phylogenetic tree.) Both of the hierarchies in α and β empirically improve model fit in the presence of heavily skewed observations (e.g. most samples are from the UK, and there is a long tail of sparsely sampled regions). We chose these model structures based on extensive cross-validation and forecasting experiments.

We place weak priors on scale parameters σ_1_, σ_2_, and σ_4_ (these denote standard deviations, the square roots of prior variance). The σ_1_ and σ_2_ priors are centered at large values to allow for wide variation in initial infection proportions across regions. The σ_4_ prior is centered around the smaller value e^−4^ ≈ 0.018 because we expect little variation of relative fitness across geographic regions a priori (some variation is expected, due to geographic variations in e.g. age distribution, behavior, or genetics as in binding affinity due HLA complex genotypes (*22*)). We fix the linear regression scale parameter σ_3_ to a small value, forcing the regression problem towards a sparse solution (i.e. we assume a priori that most observed mutations have little effect on fitness). We choose a Laplace prior on regression coefficients because it is heavier-tailed than a Normal prior, but not so heavy-tailed that the regression problem becomes multimodal (as it would for e.g. a Cauchy or Student’s t prior).

This proportional growth model differs from many forecasting models in the literature that are formulated in terms of absolute sample counts. Whereas our Multinomial likelihood allows us to model only the relative portions of lineages in each (time,region) bin, a Poisson likelihood would force us to additionally model the total number of genome samples in each (time,place) bin, a task which is less related to viral dynamics and more related to local lab capacity, political dynamics, and local calendars. We choose to model relative proportions rather than absolute counts because the relative model is robust to a number of sources of bias, including: sampling bias across regions (e.g. one region samples 1000x more than another); sampling bias over time (e.g. change in sampling rate over time); and change in absolute fitness of all lineages, in any (time, region) bin (e.g. due to changes in local policies or weather, as long as those changes affect all lineages equally). However the model is susceptible to the following sources of bias: biased sampling in any (time, region) cell (e.g. sequencing only in case of S-gene target failure); and changes in sampling bias within a single region over time (e.g. a country has a lab in only one city, then spins up a second lab in another distant city with different lineage proportions).

This model has several advantages over existing approaches. First, it provides a principled, agnostic approach that can be applied to a large dataset to identify lineages that demonstrate concerning epidemiological features. Second, by modeling the relative fitness of lineages separately across 1560 geographic regions, the model is robust to region-specific differences in non-pharmaceutical interventions and vaccination rates. Third, the hierarchical nature of the model which represents lineages as collections of mutations reflects the underlying biology and yields both strain- and lineage-specific coefficients from a single inferential approach. While the linear-additive model of mutation biology is a coarse approximation to true biology including epistasis, our hierarchical model serves as a framework to explore such models (*23, 24*) on SARS-CoV-2 genomic surveillance data.

We interpret the regression coefficients as the relative fitness based on a well-known result in population genetics <u>(Crow and and Kimura 1970)</u> that the change in genotype frequency in a large haploid population under selection follows a logistic curve, where the logistic growth rate parameter defines the relative fitness of genotypes.

#### Probabilistic Inference

The model is implemented in the Pyro probabilistic programming language (*15*) built on PyTorch (*25*). To fit an approximate joint posterior distribution over all latent variables (a space of dimension 375,909), we train a flexible reparameterized variational distribution using stochastic variational inference. Our variational approach starts by reparameterizing the model via a sequence of learnable but distribution-preserving decentering transforms (*26*) on the α and β latent variables. Reparameterizing is particularly helpful in avoiding Neal’s-funnel situations (*27*) by smoothing out the geometry of latent variables with Normal prior whose scale parameter is also a latent variable. After reparameterizing we model the posterior over all variables as a joint multivariate Normal distribution whose covariance matrix Σ is parametrized by a rank-200 matrix plus a diagonal matrix **D** with positive entries:

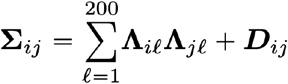

where ∧ is an unconstrained matrix of size 375,909 x 200. This low-rank multivariate Normal distribution allows the approximate posterior to capture correlated uncertainty among competing mutations each of which might explain increased fitness. This variational distribution has 75,936,525 parameters to be optimized (much larger than the number 375,909 of latent variables, but much smaller than the 375,909 ⨉ (375,909 + 1) / 2 ≅ 7 ⨉ 10^10^ parameters that would be required to represent a full-rank covariance matrix).

Variational inference is performed for 10,000 iterations with the Adam optimizer (*28*) with clipped gradients and an exponentially decreasing learning rate schedule and initial learning rates between 0.05 and 0.0025 for different parameter groups (see Figure S21). Optimization proceeds in batch-mode, i.e. without any data subsampling. We initialize model parameters to median prior values with a small amount of noise added to avoid scale parameters collapsing early in training. After inference we make predictions by drawing 1000 posterior samples. See source code for detailed optimizer and initialization configuration.

Inference and prediction on a single GPU (NVIDIA Tesla A100 with 48GB of RAM) takes about 10 minutes (compared to 14.5 hours on an 8-core CPU), which is less than the amount of time required to download and preprocess each daily snapshot of data from GISAID. The cost of fitting the model is O((TP+F)C), dominated by pointwise mathematical operations, particularly computing the softmax function on a dense array of shape T×P×C. This cost does not depend directly on the number of genetic samples, since samples are aggregated into counts y of constant shape T×P×C.

We emphasize that inference in this model is very challenging due to the large dimension of the latent space (namely 375,909), itself a consequence of the large number of regions, lineages, and mutations considered by the model (*29*). While variational inference has a number of attractive features, especially computationally, like any approximate inference scheme it comes with disadvantages. In our case the most notable disadvantage of variational inference is its tendency to yield biased posterior uncertainty estimates. Typically posterior uncertainty is underestimated, leading to credible intervals (CI) that in some cases can be unrealistically narrow. The primary parameters of interest in the PyR_0_ model are the mutation-level coefficients β_f_ and the per-lineage fitness values Σ*_f_* β*_f_ X*c*_f_*. Since the latter quantity governs the prior over β_pc_, which in turn directly feeds into the multinomial likelihood, the per-lineage fitness estimates are more-or-less tightly constrained by the observed counts y_tpc_. Consequently the posterior uncertainty of per-lineage fitness is comparatively easy to estimate and we expect variational inference to yield reasonable credible intervals for these quantities. In contrast the mutation-level coefficients β_f_ interact with correlated features X_cf_ (leading to a multi-modal posterior) and are less directly constrained by the observed counts y_tpc_. Consequently it is significantly more challenging to estimate the corresponding posterior uncertainty. In practice we obtain implausibly narrow credible intervals for these quantities and the posterior uncertainty must be interpreted with caution. Importantly, while the uncertainty estimates for β_f_ should not be taken at face value, we believe that they are still very useful for interpreting inferred model parameters, since they *can be used to rank/prioritize different hits β_f_*. In particular, while the absolute magnitudes of β_f_ uncertainty estimates are implausible, their *relative magnitudes* are representative of the amount of supporting evidence, and thus are useful for ranking. Since we consider a large number of mutations (F=2904) this information is invaluable for designing experiments for functional characterization.

#### Implementation

We implemented the PyR_0_ model using the probabilistic programming language Pyro (*15*). The model leverages PyTorch and Pyro to scale efficiently to large data sets and can therefore be applied continuously as datasets grow, completing model training and prediction with millions of viral genomes in minutes on a single GPU. We chose the Pyro framework because it cleanly separates model specification from inference customization, and scales to large models and datasets by leveraging GPUs. This flexible modeling framework allowed us to experiment with different hierarchical structures. Additionally by relying on an open source and well-tested modeling and inference framework, we minimize the risk of introducing software bugs into our analysis. The speed of inference—which took about 10 minutes on a single GPU on the full dataset of >6 million genomes—allowed quick model iteration and thorough validation on subsets of the data, including both geographic cross-validation and temporal data truncation.

#### Prediction

In Figure 3, the 95% confidence intervals in parentheses were estimated by drawing 1000 samples from the variational posterior distribution. Confirmed cases per day were estimated at the end of the training period (Jan 20 2021) by combining our model’s relative lineage portions with confirmed case count data from Johns Hopkins university. Quantities defined over our 3000 fine clusters were aggregated up to coarser PANGO lineages for reporting. To facilitate downstream use of model predictions we have provided complete tables of lineage fitness estimates (Data S1) and mutation coefficients (Data S2). These predictions have been used e.g. by Nextstrain.org to visualize our predicted mutational fitness along a phylogenetic tree (Figure S22).

#### Validation

We considered the possibility of biased submission to the GISAID database and compared results obtained from the full dataset with results obtained from disjoint subsets. For this purpose we divided the data into samples from the most heavily sampled region (Europe, with 3.3M samples) and those from the rest of the world (with 3.1M samples) (Figures S1,S11). This split is motivated by most samples originating from the UK: we widened the region around the UK until the region and its complement both had roughly equivalent statistical strength and narrow posterior estimates. We conducted two-fold cross-validation experiments for both lineages (Figure S1) and mutations (Figure S11). Additionally, in Figure S23, we show that PyR_0_ lineage-level Δ log R estimates are largely driven by regions with the largest numbers of samples and are thus robust to the manner in which under-sampled regions are organized into spatial units.

We found the full GISAID dataset to be invaluable to making accurate predictions. Using data up to July 2021, we tried restricting to either all CDC data or CDC’s randomly sampled NS3 dataset and found those subsets to result in insufficient diversity and lead to unclear results (Pearson correlation 0.49, 0.28, respectively). Using data snapshots from mid January 2022, we tried restricting to open data available in GENBANK, but found the model made implausible estimates of Omicron fitness, due to a combination of lack of geographic diversity (GENBANK has only about 1/10 as many geographic regions as we were able to extract from GISAID data, and particularly has very few samples from South Africa) and data upload latency (GISAID appeared to have ∼1 week upload latency, versus ∼1 month for GENBANK).

Our model assumes each single point mutation independently linearly contributes to change in fitness. A natural generalization is to search for groups of mutations that affect fitness. To explore this we fit a similar model of both single and pair mutations, considering only pairs that lie within the same gene. Fitting this model on data up to July 2021, we discovered no pairwise mutations stronger than the top 100 single mutations. While these experiments did not discover pairwise mutations, we believe that more sophisticated models would be able to measure epistasis, but sophistication in that area is beyond the scope of the present work.

Finally, to compare our multinomial multivariate logistic growth model to simple binomial univariate logistic growth, we compared lineage fitness estimates (Figure S24) and logistic growth curves (S25) of all but one lineage at a time, showing good agreement on the narrow selection of lineages examined by each binomial logistic fit.

### Supplemental Note 2

#### Cell culture

Cells were cultured in humidified incubators with 5% CO_2_ at 37° C, and monitored for mycoplasma contamination using the Mycoplasma Detection kit (Lonza LT07-318). HEK293 *Homo sapiens*, female, embryonic kidney cells (ATCC CRL-1573) were cultured in DMEM supplemented with 10% heat-inactivated FBS, 1 mM sodium pyruvate, 20 mM GlutaMAX, 1× MEM non-essential amino acids, and 25 mM HEPES, pH 7.2.

#### Virus production

24 hrs prior to transfection, 6 × 10^5^ HEK-293 cells were plated per well in 6 well plates. All transfections used 2.49 µg plasmid DNA with 6.25 µL TransIT LT1 transfection reagent (Mirus, Madison, WI) in 250 µL Opti-MEM (Gibco). Single-cycle HIV-1 vectors pseudotyped with SARS-CoV-2 Spike protein, either D614 or D614G, were produced by transfection of either HIV-1 pNL4-3 Δenv Δvpr luciferase reporter plasmid (pNL4-3.Luc.R-E-), or pUC57mini NL4-3 Δenv eGFP reporter plasmid, in combination with the indicated Spike expression plasmid, at a ratio of 4:1. ACE2 expression vectors were produced by transfecting cells with one of the pscALPSpuro-ACE2 plasmids, along with the HIV-1 *gag-pol* expression plasmid psPAX2, and the VSV glycoprotein expression plasmid pMD2.G (4:3:1 ratio of plasmids). 16 hrs post-transfection, culture media was changed. Viral supernatant was harvested 48 hours after media change, passed through a 0.45 µm filter, and stored at 4°C. TMPRSS2 expression transfer vector was produced similarly but with pscALPSblasti-TMPRSS2.

#### Generation of cell lines expressing ACE2 and TMPRSS2

2.5 x 10^5^ HEK-293 cells were plated per well in a 12 well plate. The next day cells were transduced with 250 uL of supernatant containing TMPRSS2-encoding lentivirus for 16 hr at 37°C, after which fresh media was added to cells. 48 hrs after transduction cells were replated and selected with blasticidin (InvivoGen, catalogue #ant-bl-1) at 10 ug/ml. After selection, cells were transduced similarly with supernatant containing ACE2-encoding lentivirus and selected with 1 ug/mL of puromycin (InvivoGen, San Diego, CA, catalogue #ant-pr-1).

#### Virus Infectivity Assays

16 hours prior to transduction, adherent cells were seeded in 96 well plates. HEK-293 cells were plated at 5 x 10^4^ cells per well. Cells were incubated in virus-containing media for 16 hrs at 37°C when fresh medium was added to cells. 48 to 72 hours after transduction cells were assessed for luciferase activity. Cells transduced with luciferase expressing virus were assessed using Promega Steady-Glo system (Promega Madison, WI). GraphPad Prism 8.4.3 was used to analyze the infectivity data using a ratio paired t test. In these experiments, all values shown are the mean with standard deviation, with the actual calculated two-tailed *P* value indicated.

### Supplemental Note 3

We include here an extended discussion of high-scoring mutations.

#### Relation to other viruses

The concentration of putative transmission-promoting substitutions in N at positions 160-210 is remarkable, but is supported by a similar observation in Ebola virus(*30*), and recent data for SARS-CoV-2 showing mutations in that region increase the efficiency of viral packaging(*31*), validating some of the model’s most unexpected predictions and supporting its ability to identify novel biology.

#### Potential functional roles of mutations within ORF1

Our model highlighted mutations within the ORF1 non-structural proteins (nsps) whose functions are not fully understood (e.g. Table S3). We found two predominant clusters within ORF1a: one in the C-terminal ∼120 amino acids of nsp4 and the other within the N-terminal ∼160 amino acids of nsp6 (Figure S13C). Nsp4 and nsp6 are both membrane-anchored proteins with roles in assembly and concentration of the viral replication and transcription complex (RTC) machinery within double-membrane vesicles (*32*). Amino acid substitutions in these regions, combined with transmission-associated mutations identified within additional RTC-associated nsps (e.g., nsp12-16, Figure S13D), may therefore affect the kinetics of replication and gene expression, resulting in higher virus yields from infected cells. Nsp2, a rapidly evolving accessory protein (*33*)(*34*)(*35*) whose proposed function in disrupting host cell signaling (*36*) and viral mRNA translation initiation (*37*) remains obscure, harbored many additional mutations associated with higher fitness (Figure S13C).

The ORF1a-ORF1b polyprotein is processed into 16 non-structural proteins by two viral proteases: a papain-like protease (nsp3) and 3C-like protease (nsp5). Multiple transmission-associated mutations were found within the protease coding regions (e.g., ORF1a:V1750A, ORF1a:P3395H). Most of the amino acid substitutions identified by our model were outside of the domains containing catalytic residues for nsp3 (C1674, H1835, D1849) or nsp5 (H3304, C3408) (*38*)(*39*). However, the potential effects of these mutations on protease architecture and activity warrant further experimentation. A few of the top mutations from our model (e.g., ORF1a:T3255I, ORF1a:A3571V) are positioned adjacent to nsp cleavage sites, potentially influencing local structures and kinetics of polyprotein processing by nsp3 and nsp5 (Figure S13C-D).

Multiple highly-ranked mutations are distributed across the replication and transcription-associated nsps in ORF1b (Figure S13D). The P314L (P323L) mutation in nsp12 – the viral RNA-dependent RNA polymerase (RdRP) – emerged early during the pandemic and became established in circulating lineages alongside S D614G (*6*). A later variant at this site (P314F) was also highly ranked in our list. Additional mutations in nsp12 can be found within the canonical fingers (D445A, V631I, D514N, G662S), palm (M592I, H604Y, T701I, C721R, S763F), and thumb (L820F, L829I, D870N) subdomains of the RdRP conserved catalytic fold (Figure S15). The functional effects of these mutations on polymerase processivity and fidelity remain to be investigated. A structural model of the SARS-CoV-2 polymerase complex has been resolved (*40*)(*41*), and contains a single subunit of nsp12, two subunits of the nsp13 helicase, and additional RdRP cofactor proteins (nsp7, 8, and 9). The ORF1b P314 residue is located at the interaction interface between nsp12 and a single subunit of nsp8. Moreover, several of the top mutations from our dataset ORF1b (e.g., P1000L, P1001S, Q1011H) are harbored within the nsp13 N-terminal zinc-binding domain that directly interacts with nsp8 (*42*). These findings implicate transmission-associated mutations within the SARS-CoV-2 RNA synthesis machinery in altering the stability of the replication complex, possibly via interactions with nsp8.

Nsp14 is a dual-functional enzyme with N-terminal 3’-to-5’ exonuclease (ExoN) and C-terminal guanine-N7 methyltransferase (N7-MTase) activities (*43*)(*44*) and is a core component of the coronavirus RNA proofreading complex. Nsp14 is uniquely responsible for excision of mismatched bases from the nascent RNA and methylation of the viral mRNA cap structure. Two mutational hotspots in nsp14 map to discrete regions in the ExoN (e.g., T1540I, I1566V) and N7-MTase (e.g., D1848Y, P1936H) domains. The functional consequences of these clusters of transmission-associated mutations on mRNA synthesis and genome replication remain unknown.

### Supplementary Figures

**Figure S1.**
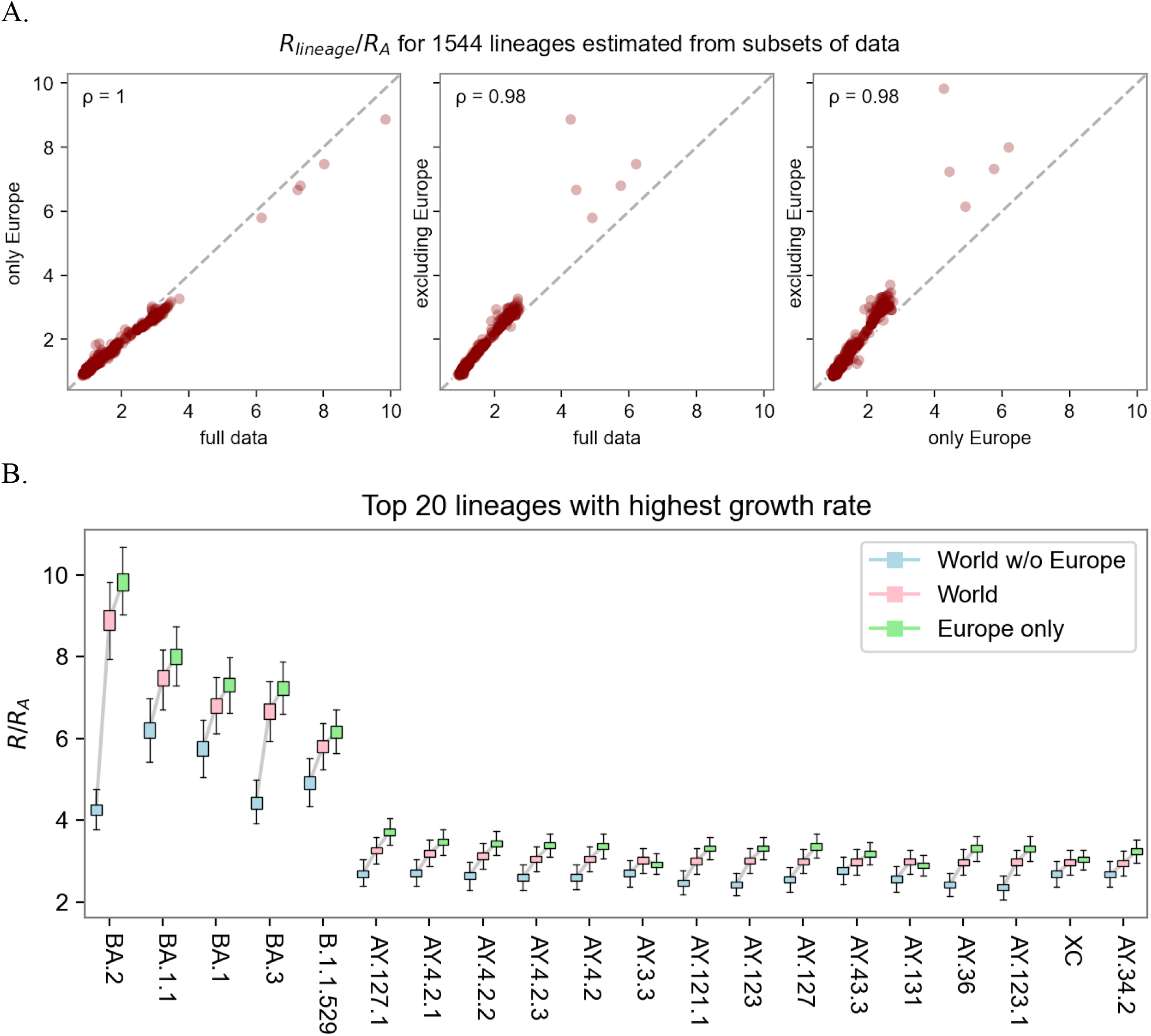
**A**. Sensitivity of lineage fitness estimates to data subset. We depict the relative fitness of all lineages as estimated by either the full data or two disjoint geographic subsets (within Europe and outside Europe). High Pearson correlation (ρ) suggests estimates are largely insensitive to data subset. **B**. Estimates of fold increases in fitness for the top 20 lineages. Sensitivity analysis shows consistency across estimates from subsets of the data in different geographic regions.

**Figure S2.**
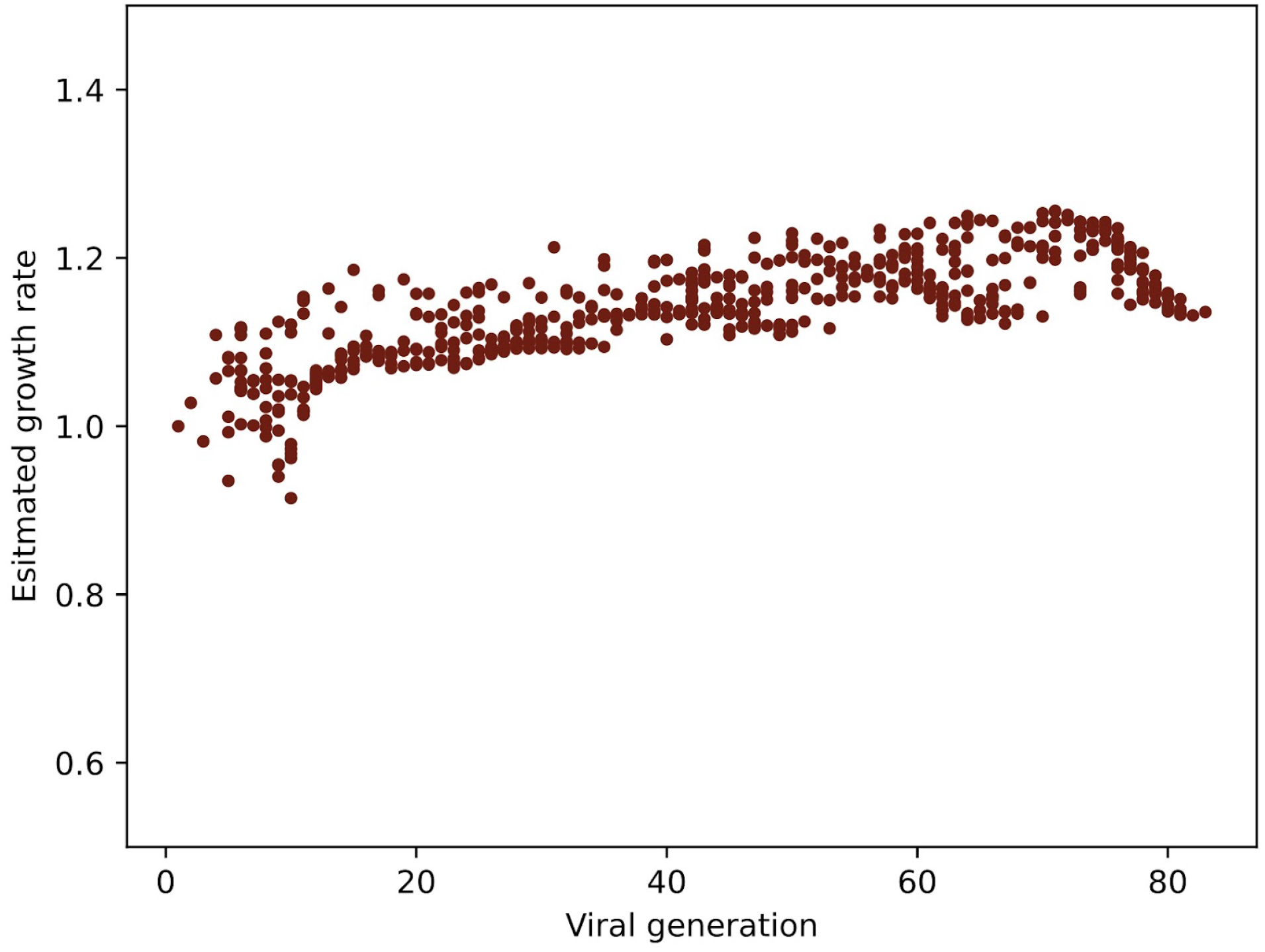
Simulation study assessing bias. Distribution of inferred fitness of new lineages as a function of time, for a simulated neutrally evolving viral population. The most successful subclades of each generation are designated as new lineages, leading to a trend toward higher estimated fitness even though all lineages are equally transmissible.

**Figure S3.**
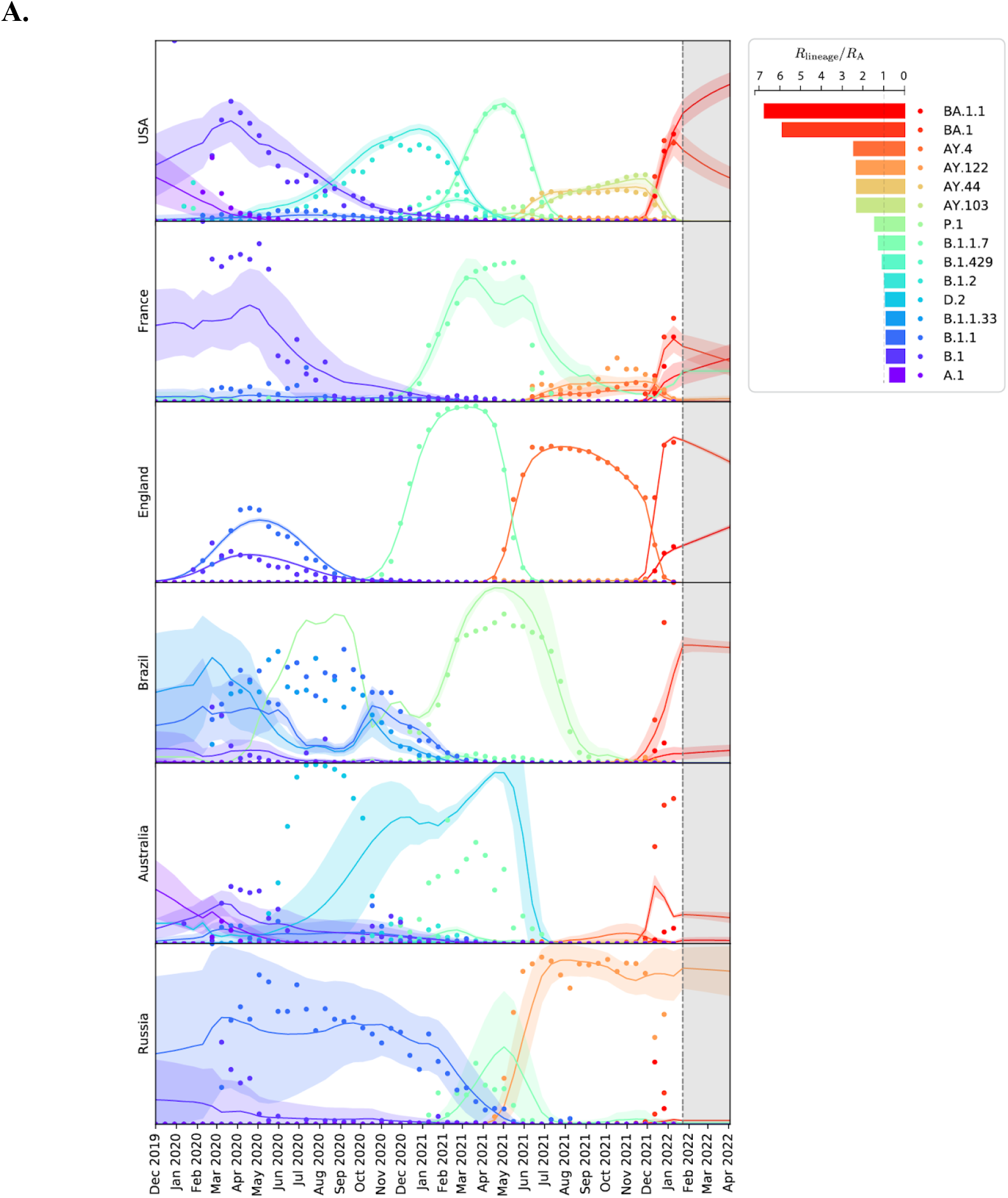
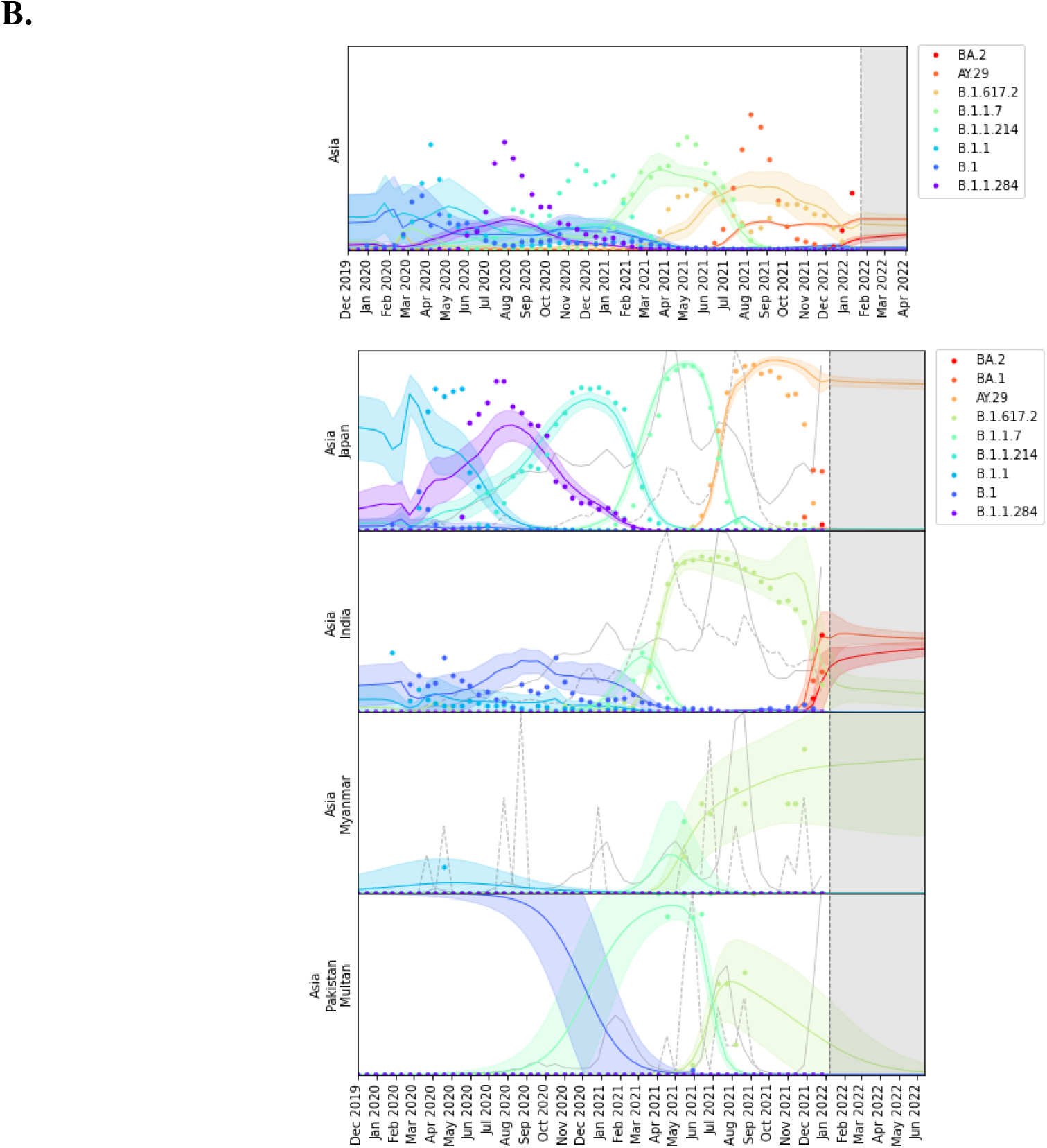
**A.** Regional fits and forecasts for USA, France, England, Brazil, Australia and Russia. Solid circles at the beginning of each two week time interval denote observed lineage proportions on a [0, 1] scale for the top 20 lineages. Solid curves and 95% confidence bands denote model predictions and three-month forecasts. Each of the six (aggregate) regions is made up of multiple subregions. The behavior of each SARS-Cov-2 cluster in each subregion is represented by only two numbers in the model: a slope and an intercept. The complex model fit results from the multivariate logistic function applied jointly to multiple competing trends, which are then aggregated over subregions and multiple clusters per lineage. England shows clear waves of dominance: B.1.1, B.1.177, B.1.1.7 (Alpha), AY.4 (Delta), and finally BA.1 (Omicron), with the latter currently being overtaken by BA.1.1 (also Omicron). Massachusetts and Brazil both start with very low sampling rates early in the pandemic. The legend reports the estimated fitness for the top 15 lineages. **B.** Region-specific fits for several regions in Asia, demonstrating better fits in regions with high sampling (Japan, India), and degraded fits in regions with low sampling (Myanmar, Pakistan).

**Figure S4.**
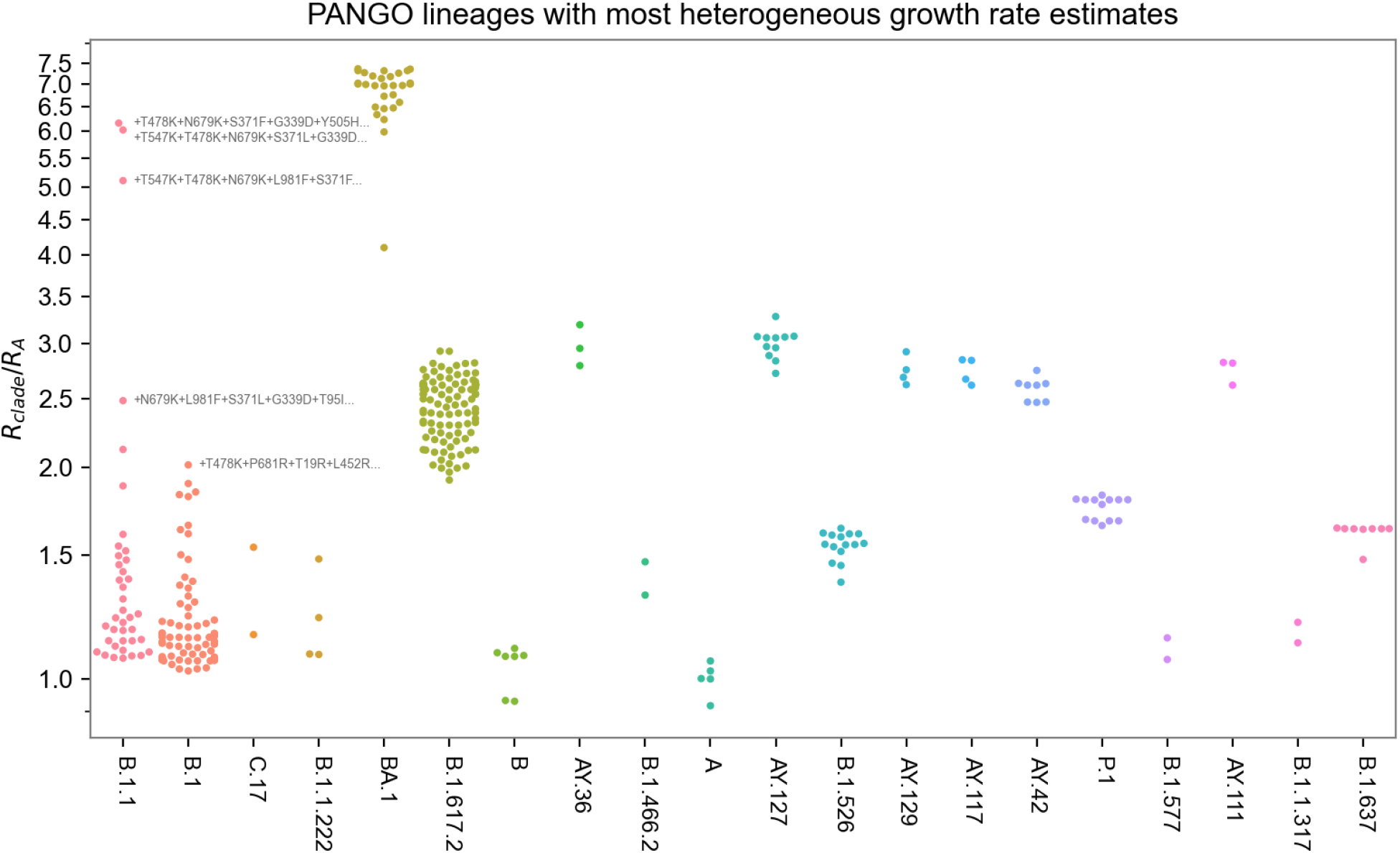
Heterogeneity of PANGO lineages. We hypothesized that the PANGO lineage clustering conflated viruses with distinct growth rate, e.g. B.1.1 exhibits two peaks in relative abundance in England, contrary to our multivariate logistic model. To test this hypothesis we refined 1544 PANGO lineages into 3000 finer clusters and estimated each cluster’s growth rate. As shown in the figure some PANGO lineages include clusters with estimated fitness differing by more than a factor of 6, including the B.1.1 lineage. This heterogeneity is also reflected in the temporal structure: for example, the three B.1.1 clusters with the largest growth rate emerged in December 2021 and January 2022, whereas the majority of B.1.1 clusters emerged in the twelve months leading up to April 2021. The top four clusters in B.1.1 and the top cluster in B.1 are labeled by their top 5 fitness-increasing mutations to the S gene, relative to the PANGO lineage’s basal sequence.

**Figure S5.**
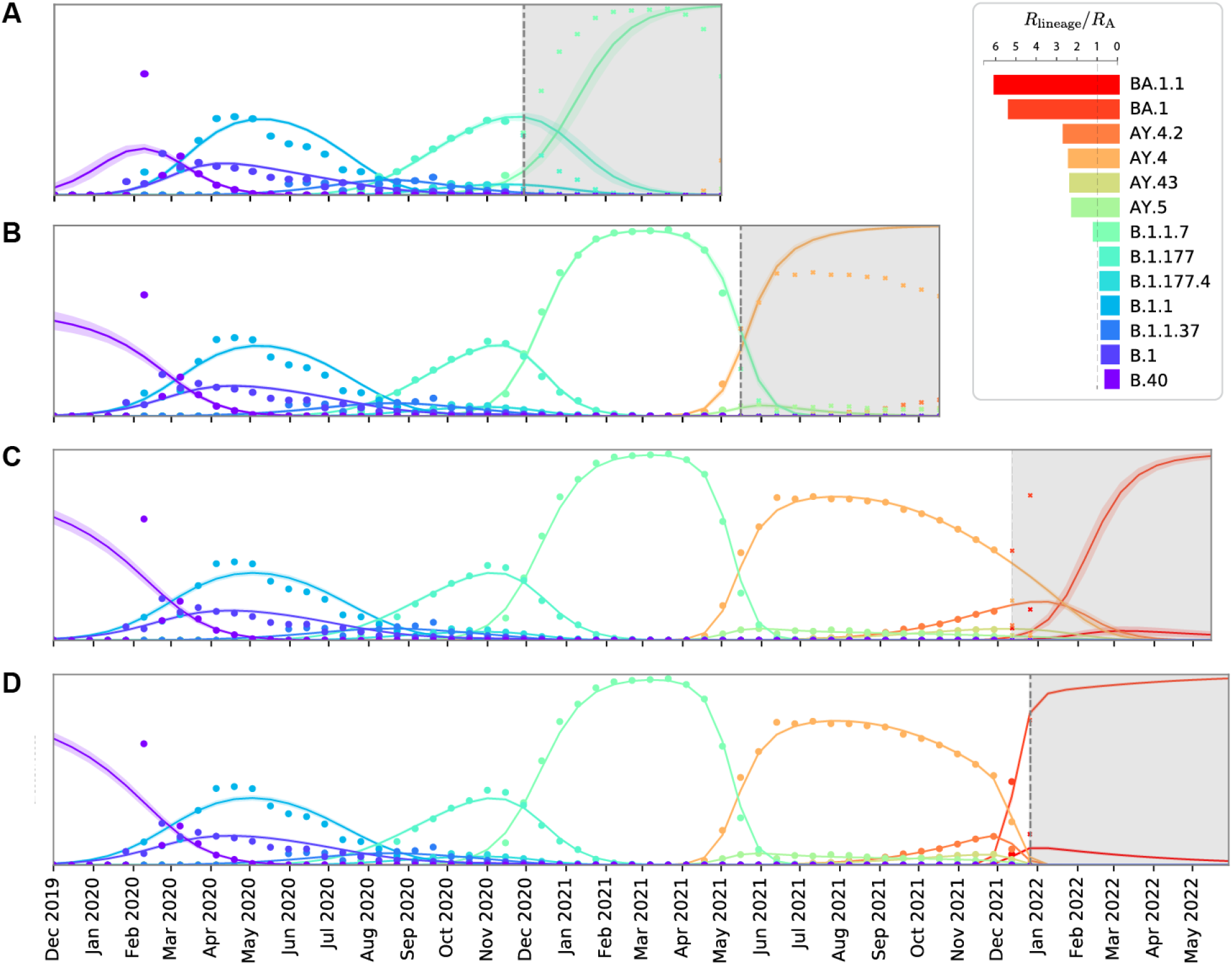
Forecasts in England with time-truncated input data. (A) Prediction for rise of B.1.1.7 using data through late November 2020 (solid circles at the beginning of each two-week time interval). (B) Prediction for rise of AY.4 using data through early May 2021. (C) Prediction for rise of BA.1 using data through mid December 2021, and (D) late December 2021. Future data points, not used during the model training, are shown in crosses. The legend reports lineage fitness estimates based on all available data.

**Figure S6.**
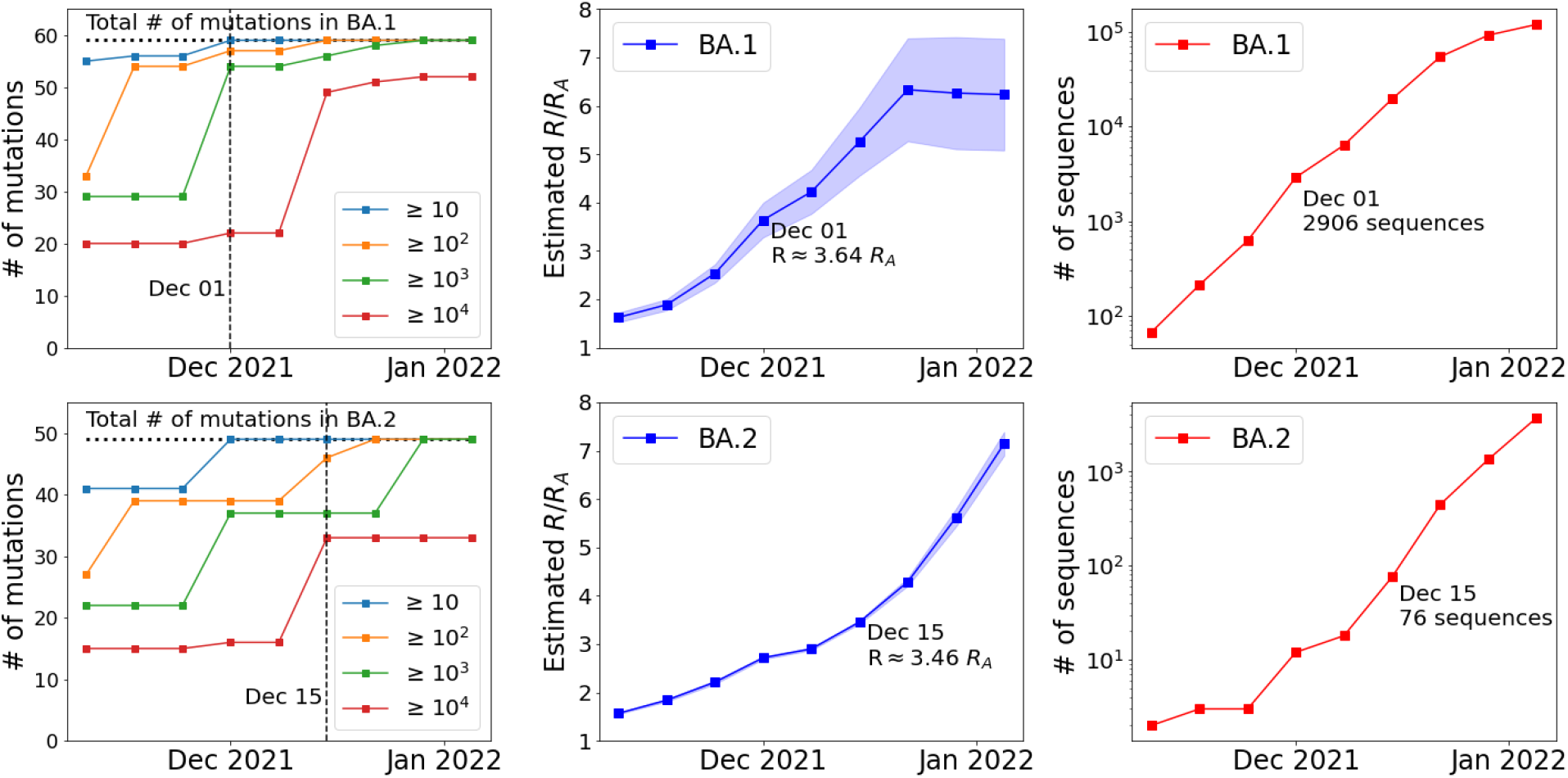
We depict the ability of PyR_0_ to predict the fitness of Omicron sublineages BA.1 and BA.2 as the number of sequenced genomes increased throughout the last two months of 2021. PyR_0_ predicted that BA.1 (respectively, BA.2) was substantially more fit than Delta by December 1^st^ (15^th^) 2021, by which time 2906 (76) genomic sequences had been collected. The substantial heterogeneity of the BA.1 sublineage is reflected in the uncertainty in BA.1 R estimates; this heterogeneity also helps explain why PyR_0_ required more sequences to identify the elevated fitness of BA.1 as compared to the case of BA.2. **Left:** The number of amino acid mutations that make up BA.1 and BA.2 that had been observed in at least 10/10^2^/10^3^/10^4^ sequences by the given date. **Middle:** Estimates of R/R_A_ using sequences collected by the given date. **Right:** The total number of BA.1 and BA.2 sequences collected by the given date.

**Figure S7:**
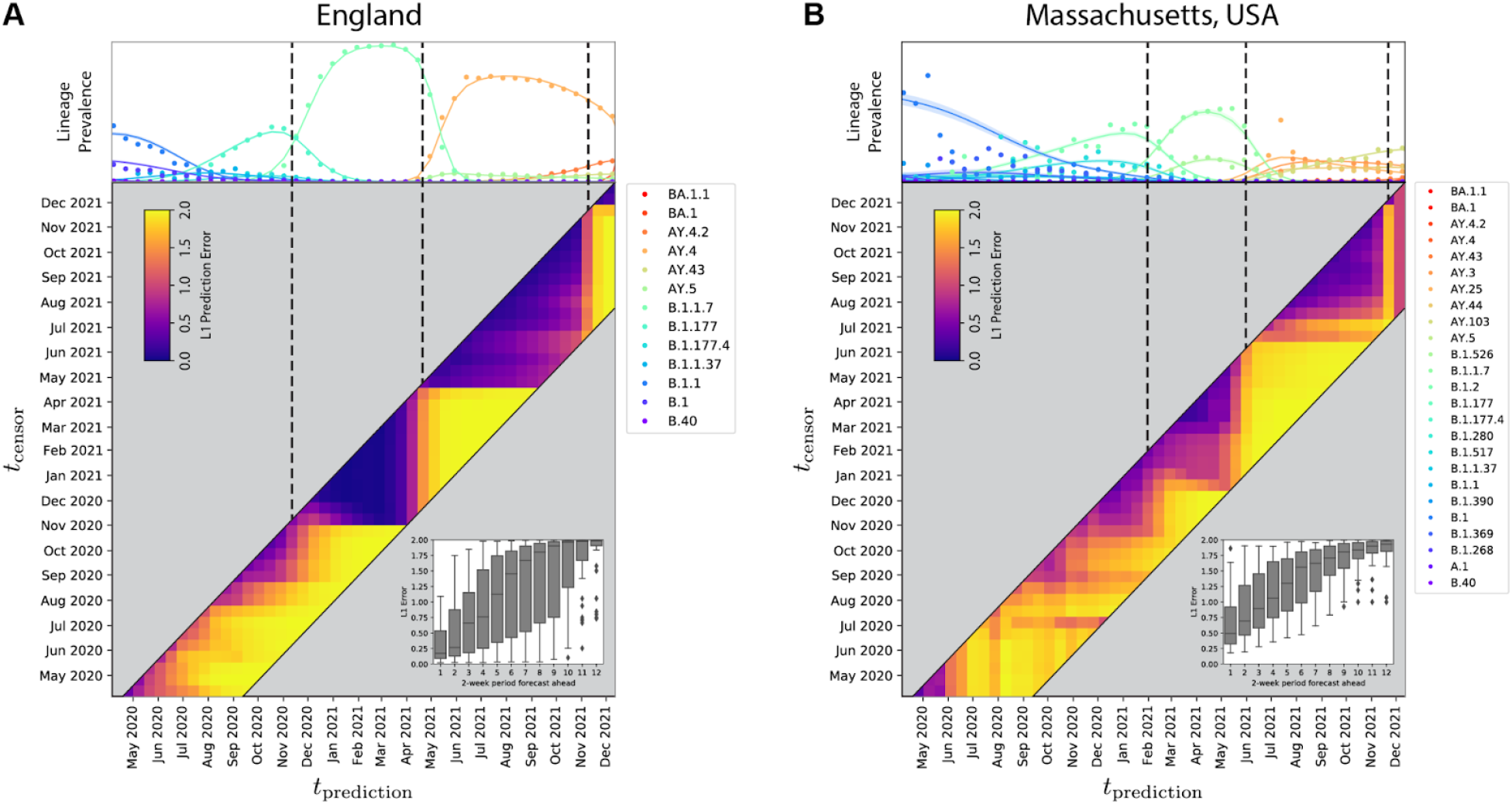
Forecasting evaluation based on independently trained models at 45 time points during the pandemic, t_censor_, and predicting at time t_predicted_ up to 12 two-week periods into the future. The results are shown for (A) England and (B) Massachusetts, USA. The top panels are as in Figure S3, heatmaps depict the prediction L1 error, and the inset bar plots depict the aggregated prediction errors over all periods. Note the rapid increase in error as new fit lineages emerge in a region (vertical dashed lines provided as a guide to the eye), followed by rapid recovery and stabilization of forecasting accuracy within only a single period, highlighting the predictive value of PyR_0_ for detecting variants of concern. Refer to Table S1 for tabulated forecasting accuracy figures in several other regions.

**Figure S8.**
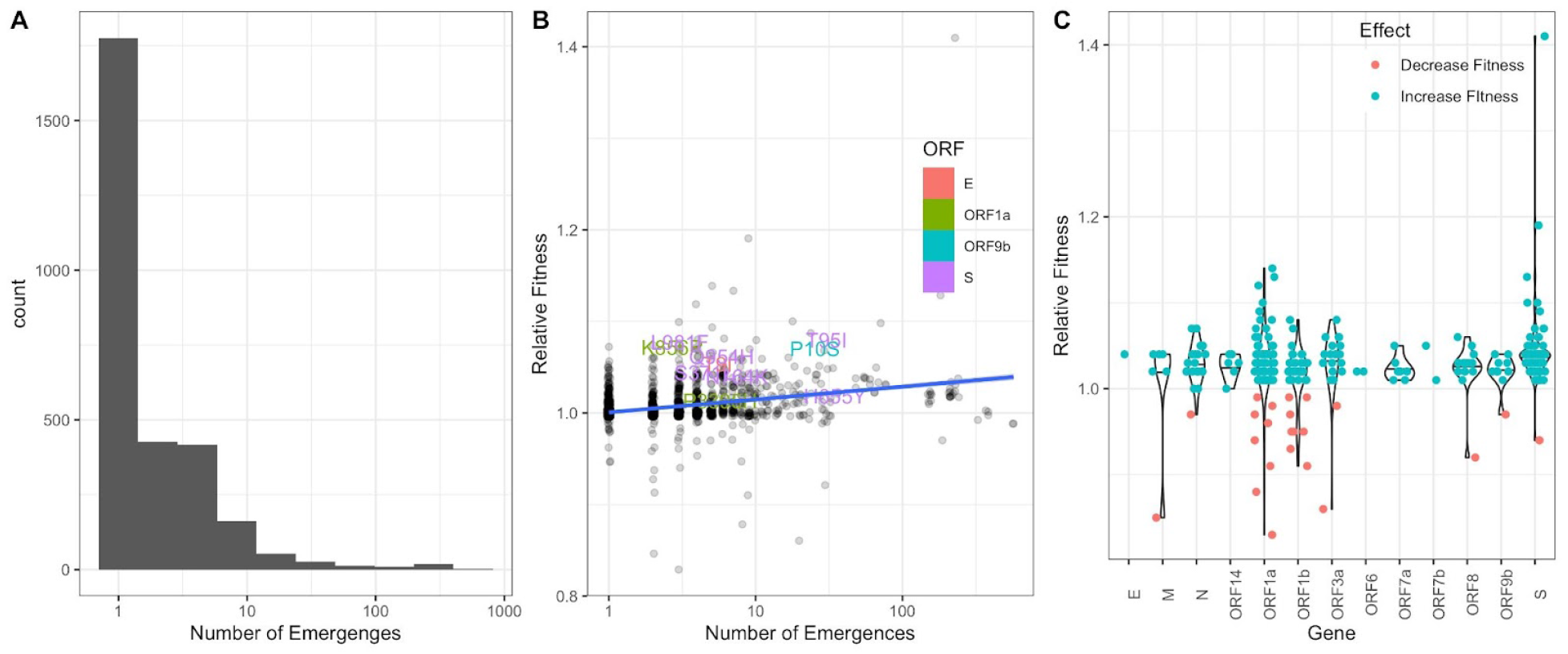
**A.** Histogram of the number of independent emergences across all observed mutations. A mutation was considered to have emerged independently if it was present in a lineage but not in its parent. **B.** Scatterplot of the fold-change in fitness versus the number of independent emergences. The top 10 ranked mutations are labeled, colored by ORF. Linear regression with standard error for the slope given as shaded area. **C.** Violin plots of fold-change in fitness for mutation, grouped by gene. The top 10% most statistically significant mutations are shown (where significance is determined by z-score of the approximate variational posterior).

**Figure S9.**
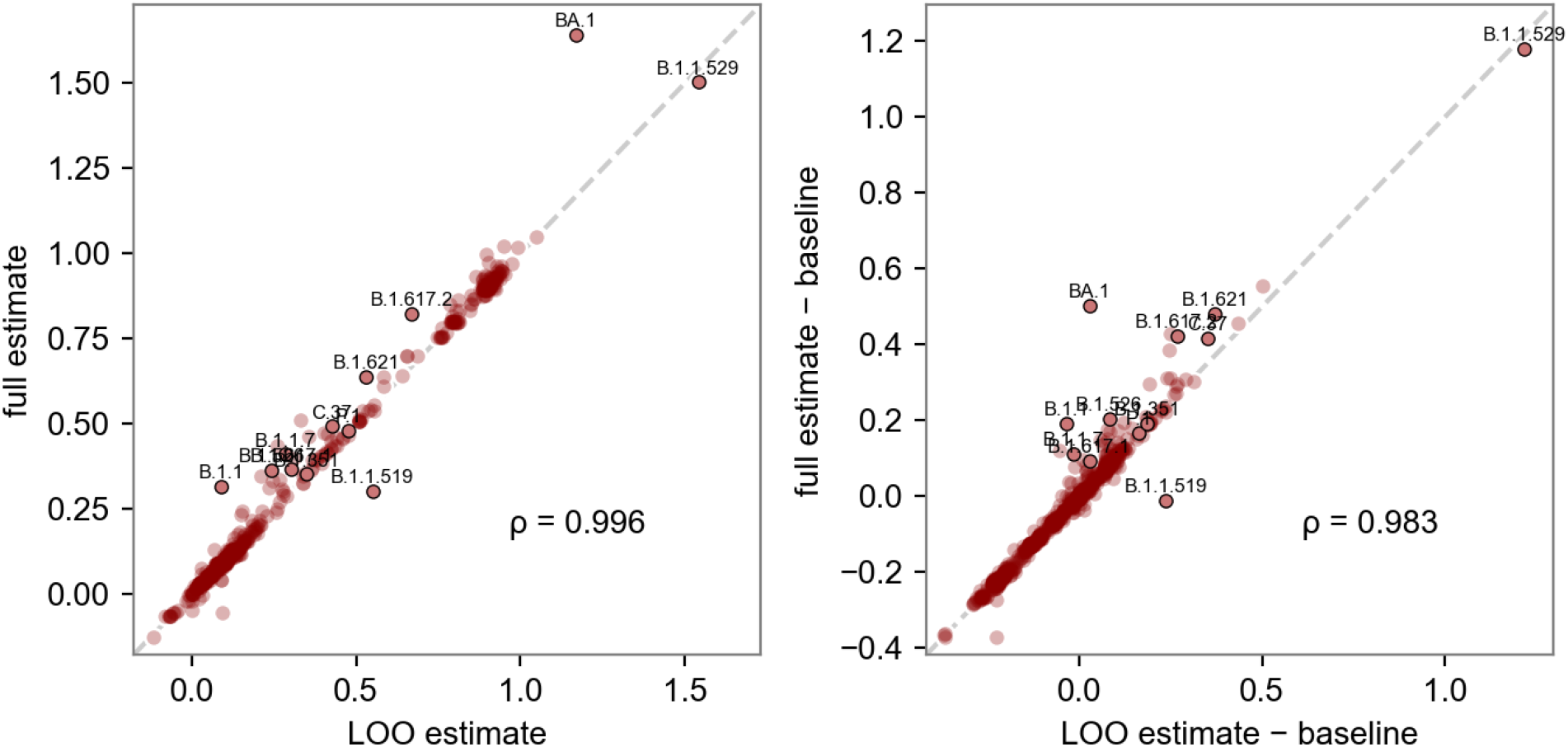
Accuracy of predicted fitness based solely on mutation content. **Left:** estimated log(R/R_A_) for each lineage based on the full set of GISAID samples (y axis), and on the leave-one-out subset with each lineage’s subclade removed and the fitness estimated from the mutations present in the lineage (x axis). **Right:** the same quantities but relative to a baseline estimator in which each child lineage’s fitness is the same as that of its parent lineage. If a mutation is entirely removed from the LOO dataset, then the corresponding mutation coefficient is estimated as zero. The evaluation metrics are Pearson correlation (⍴) and mean absolute error (MAE). The MAE of the leave-one-out estimator is 0.001, more than 100x smaller than the MAE of the baseline estimator (0.129). Both panels highlight the CDC’s variants of concern and variants of interest. The lineages selected for testing are those with at least 100 samples and with the largest deviations from their parent, i.e. where the baseline estimator performs worst. Note that the fitness of child lineages can deviate substantially from that of the parent, e.g. BA.1 is surprisingly higher fitness than its parent B.1.1.529.

**Figure S10.**
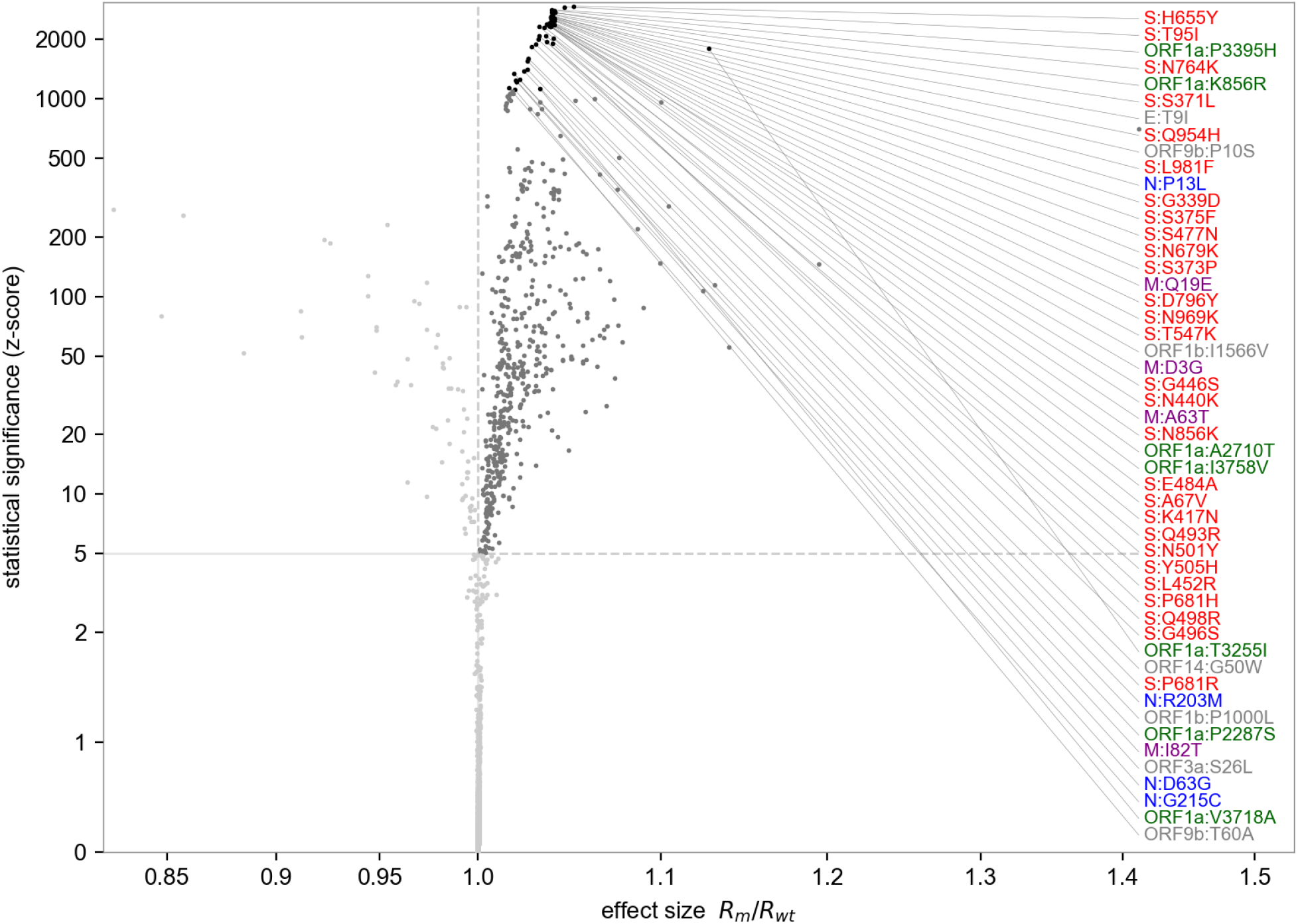
Volcano plot highlighting the most statistically significant mutations linked to increased fitness. The x-axis depicts the effect size as a ratio of estimated fitness of lineages with-versus-without each mutation. The y-axis depicts z-score from the approximate variational posterior as a proxy for statistical significance. The top 50 most statistically significant mutations are labeled, colored by gene. The 540 growth-associated mutations with z-score greater than 5 are shaded dark gray.

**Figure S11.**
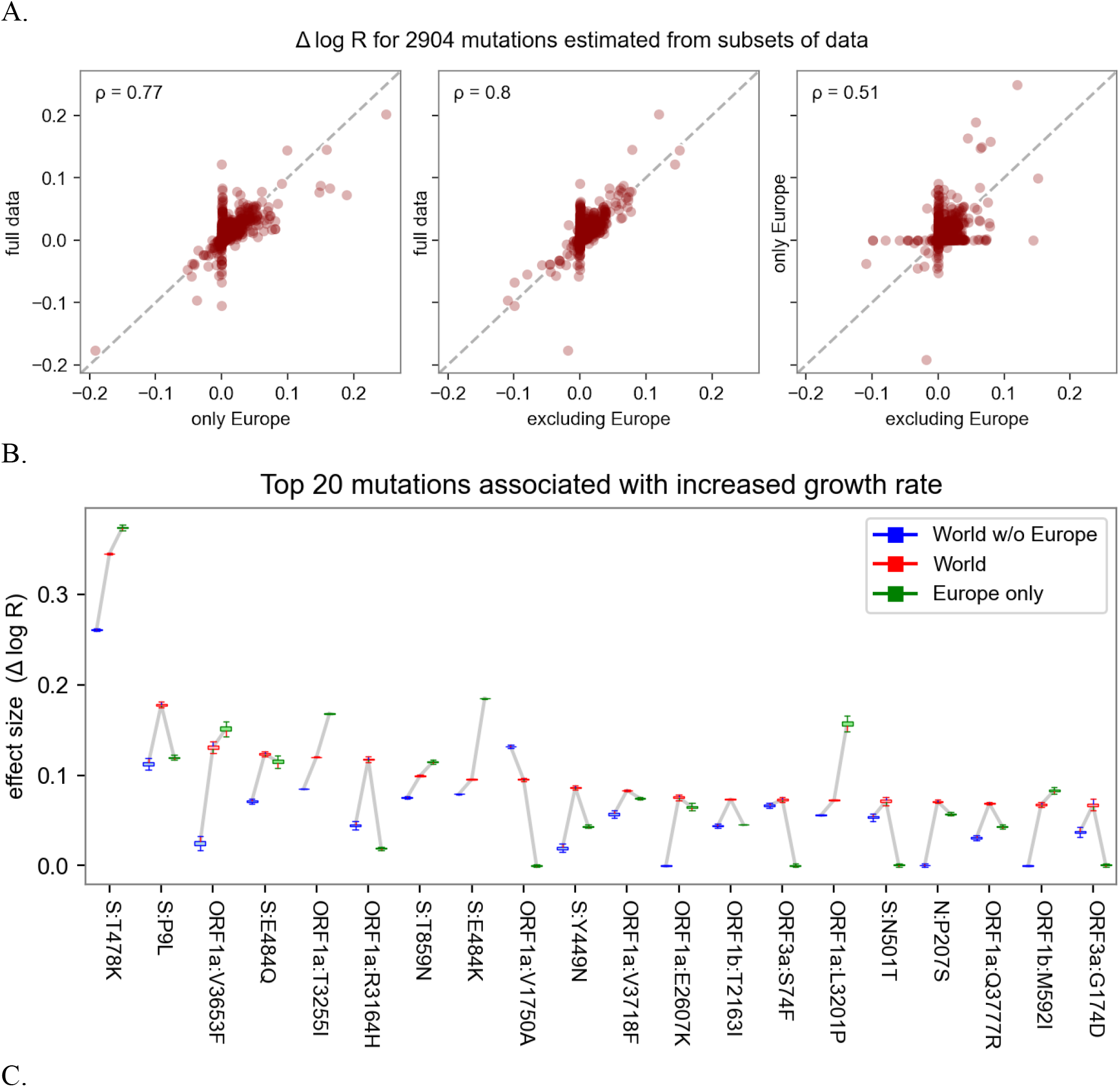
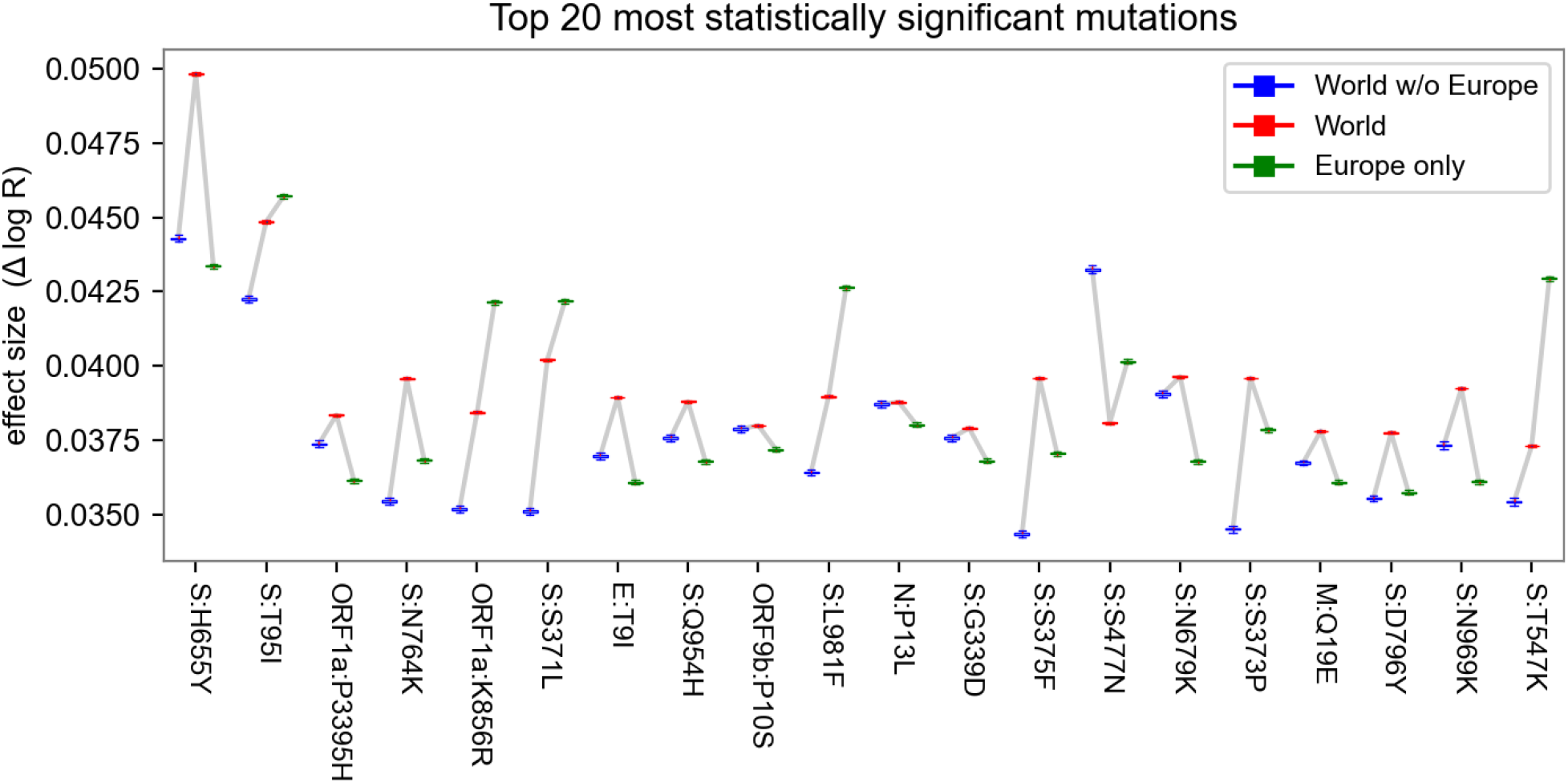
Sensitivity of mutation estimates. **A.** Scatterplot of the mutation coefficients on the full model and geographic subsets, with Pearson correlation (ρ) as shown. **B.** Box-and-whisker plot depicting estimated growth rates with corresponding uncertainties for the 20 lineages with highest growth rate (effect size) across geographic subsets. **C.** Same as B but with the top 20 lineages sorted by statistical significance (z-score). Note that in B, the World estimates (center) tend to be higher than subsets (left and right) only because the ranked selection is based on those estimates.

**Figure S12.**
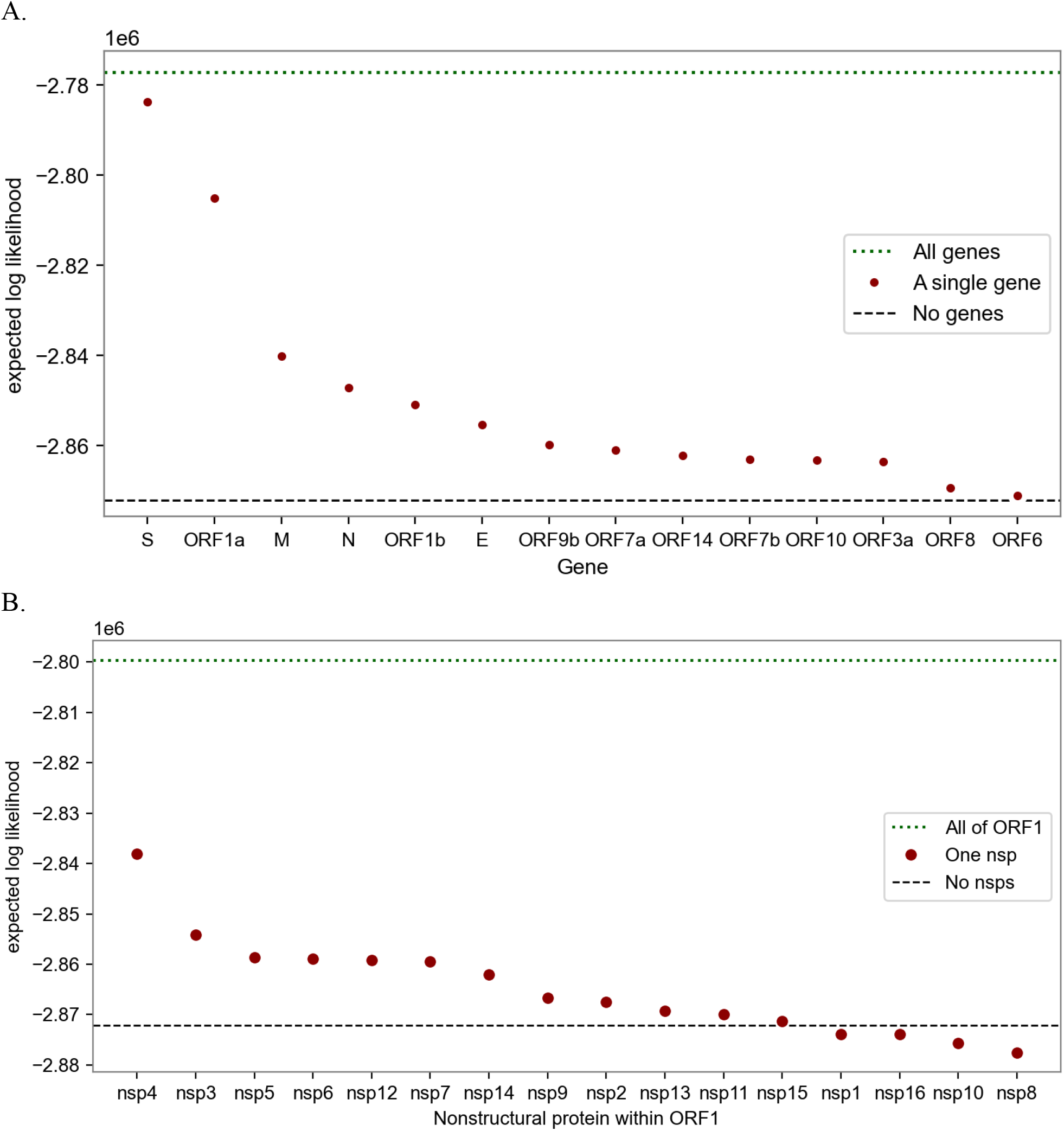
Information content of different subsets of the SARS-CoV-2 genome in explaining fitness. The metric is the expected log likelihood. The dotted line at the top shows the performance of the full model that regresses against all genes (A) or against all of ORF1 (B). The circles show estimators based on only single genes (A) or single nonstructural proteins (B). The most informative genes are S, ORF1a, M, N, and ORF1b; within ORF1 the most informative nsps are nsp4, 3, 5, 6, 12 and 7. The bottom dashed lines show the performance of a naive estimator that ignores genetic information, effectively estimating each lineage’s growth rate in each region independent of growth rate estimates in other regions.

**Figure S13.**
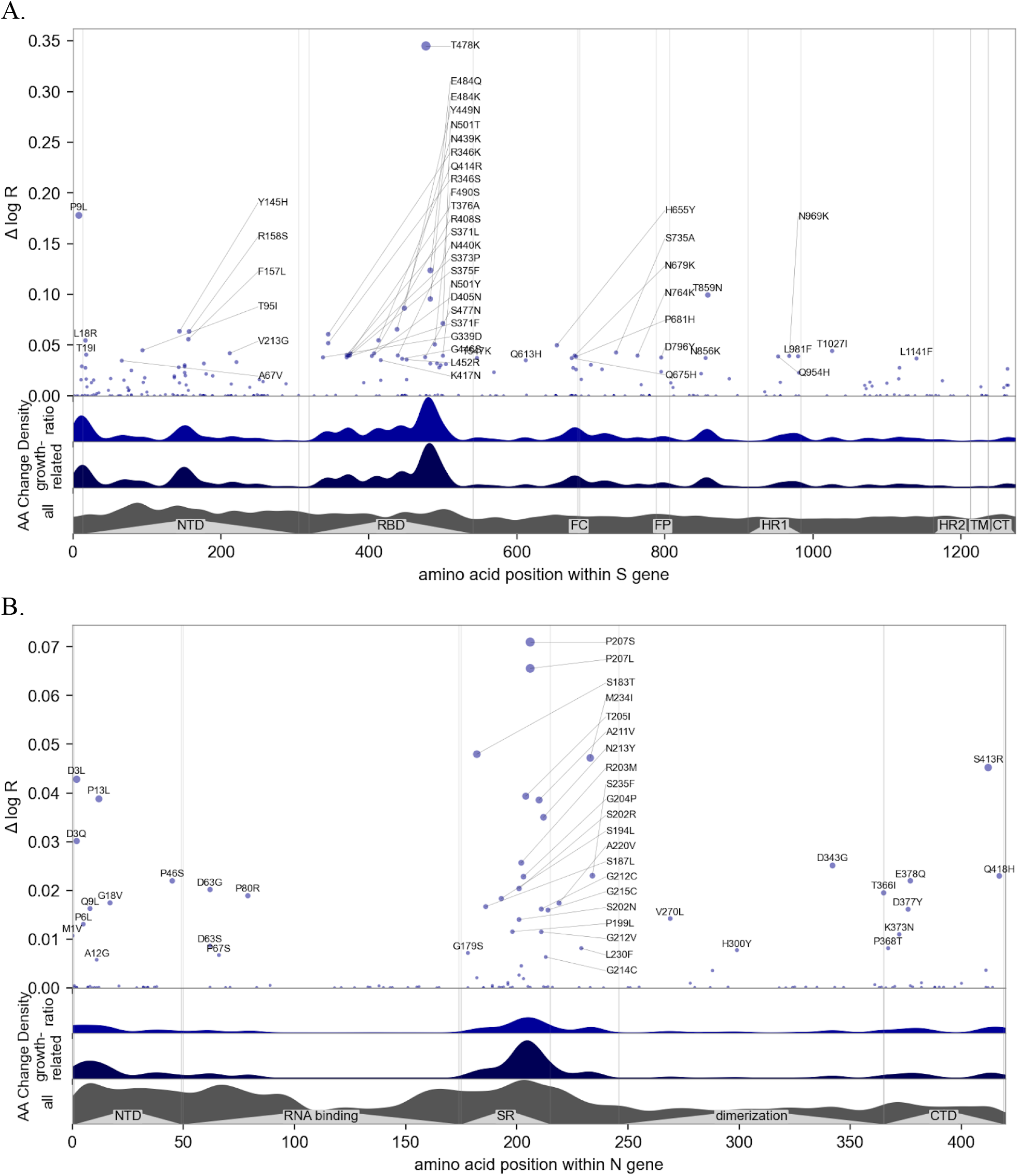
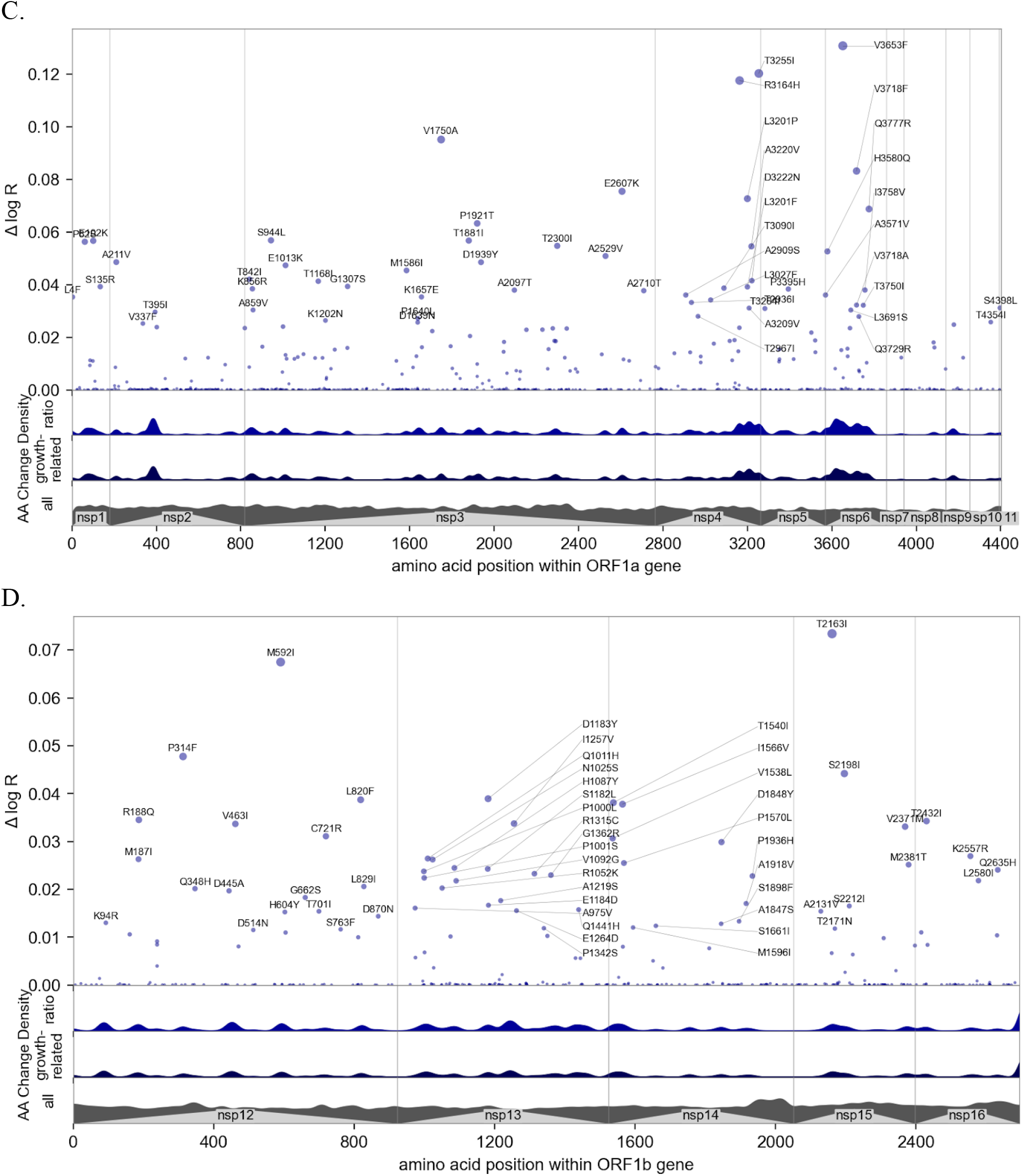
Manhattan plot details of the four most informative genes. See Figure 3 for a whole-genome view and Figure S12 for ranking by information. **A.** View of the 1237 amino acids of the S protein, annotated by structure (45); many mutations occur in the N-terminal domain (NTD), receptor-binding domain (RBD), and furin cleavage (FC) site. Regions containing the fusion peptide (FP), heptad repeat (HR) 1 and 2, transmembrane domain (TM), and C-terminal domain (CTD) are annotated. **B.** View of the 419 amino acids of the nucleocapsid (N) protein domains, annotated by structure (46); many mutations occur in the serine–arginine rich region (SR), identified by (47) as immunogenic. **C.** View of the ORF1a polyprotein, including 11 non-structural proteins (nsps). **D.** View of the ORF1b polyprotein, including nsp12-16; note the amino acid positions do not account for 9 additional residues at the N-terminus of nsp12 (RNA polymerase) resulting from the -1 ribosomal frameshift.

**Figure S14:**
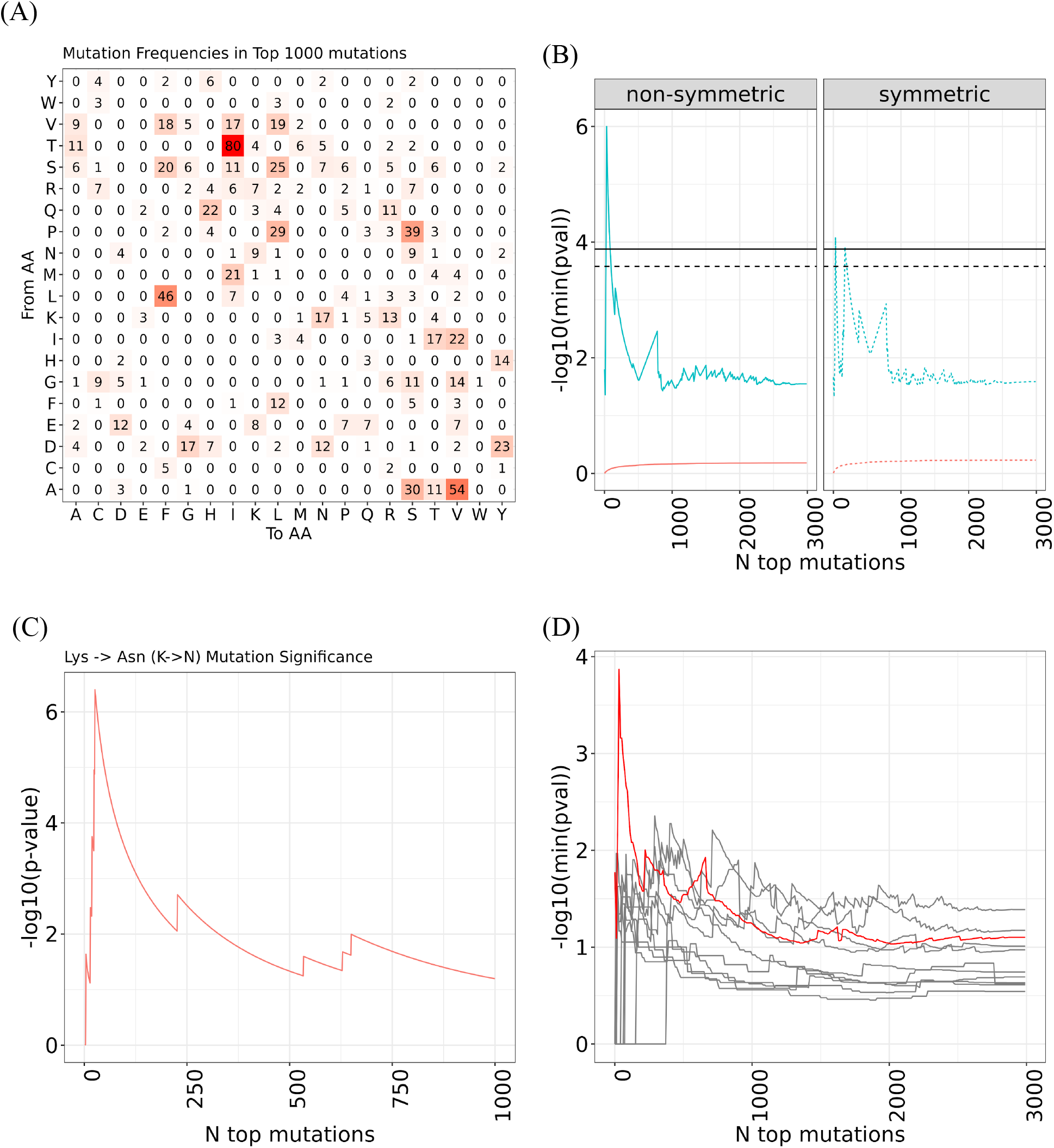
Enrichment analysis of amino acid changes among top-ranked mutations **A.** Mutation frequency in top 1000 most significant mutations (as ranked by posterior mean/stddev) **B.** Leading set enrichment analysis of most significant mutations predicted by the model for non-symmetrical (e.g. A->V) (left) and symmetrical (e.g. A<-<V) (right) amino acid changes. The blue curve depicts the most significant p-value obtained for different top-N mutation cutoff values across all amino acid changes, while the red curve depicts the mean p-value. **C.** Further examination reveals that top mutations are enriched in K to N changes in the S gene. **D.** No other genes (gray) other than S (in red) show significant enrichment.

**Figure S15.**
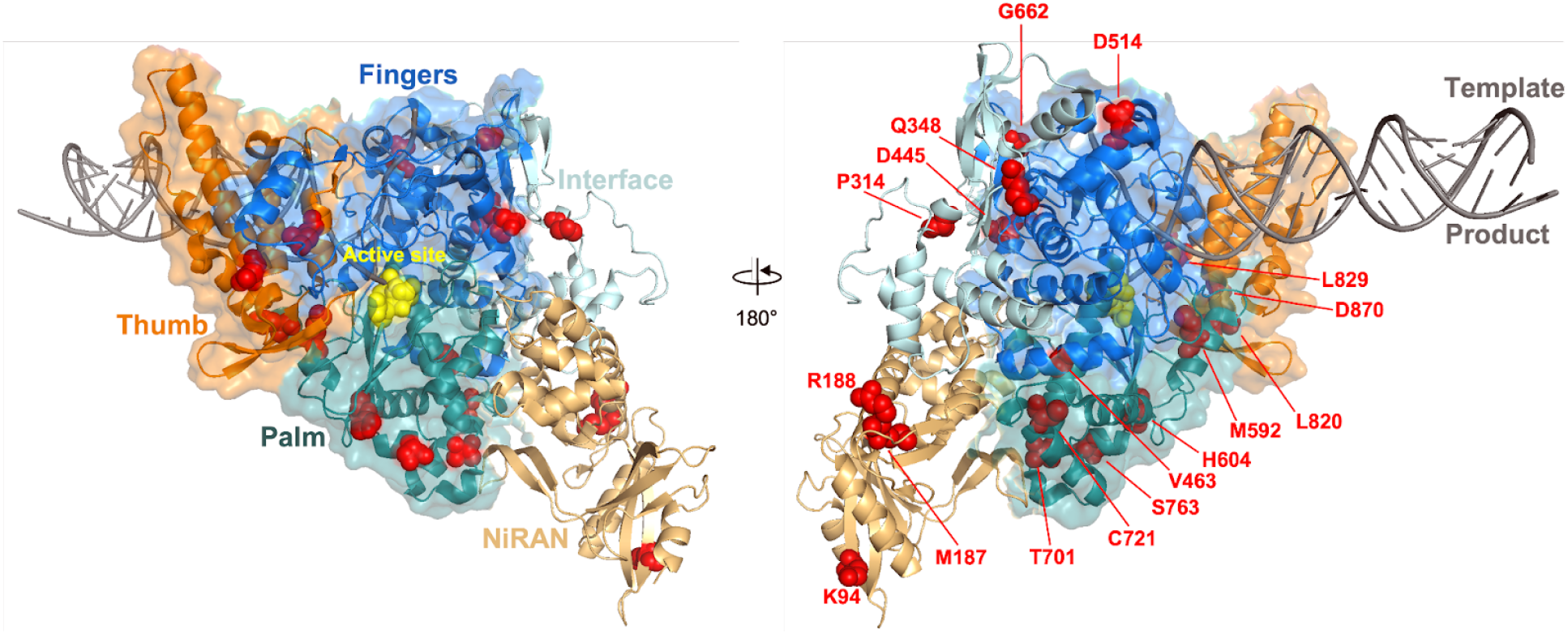
Top-ranked mutations in the viral RNA-dependent RNA polymerase (RdRP, nsp12, PDB: 7CYQ). Amino acid positions corresponding to top mutations are shown as red spheres. The catalytic site residues 750-SDD-752 are highlighted as yellow spheres. The coronavirus-specific domains (NiRAN, Interface) are shown as cartoon structures. The conserved RdRP domains (Fingers, Palm, Thumb) are shown as cartoon and surface filling structures

**Figure S16.**
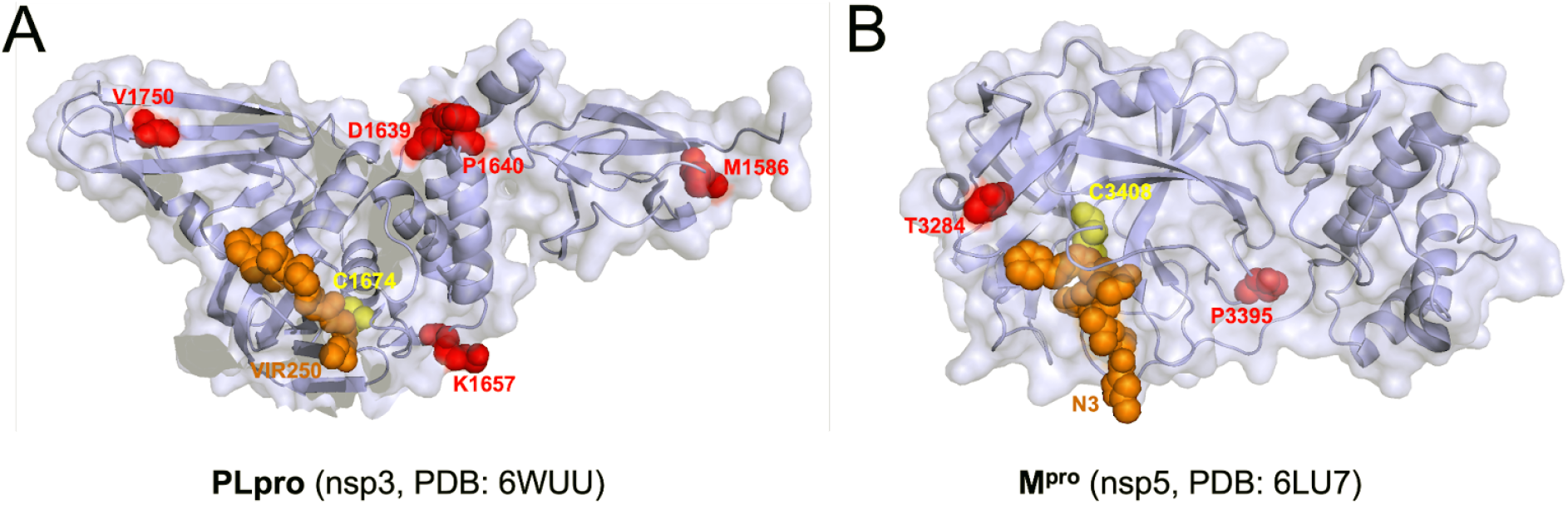
Top-ranked mutations in the two viral proteases, PLpro (A) and M^pro^ (B). Both protease structures are shown in light blue. Amino acid positions corresponding to top mutations are shown as red spheres. The catalytic cysteine residues for each are shown as yellow spheres. Two active-site inhibitors, VIR250 and N3, are shown as orange spheres.

**Figure S17.**
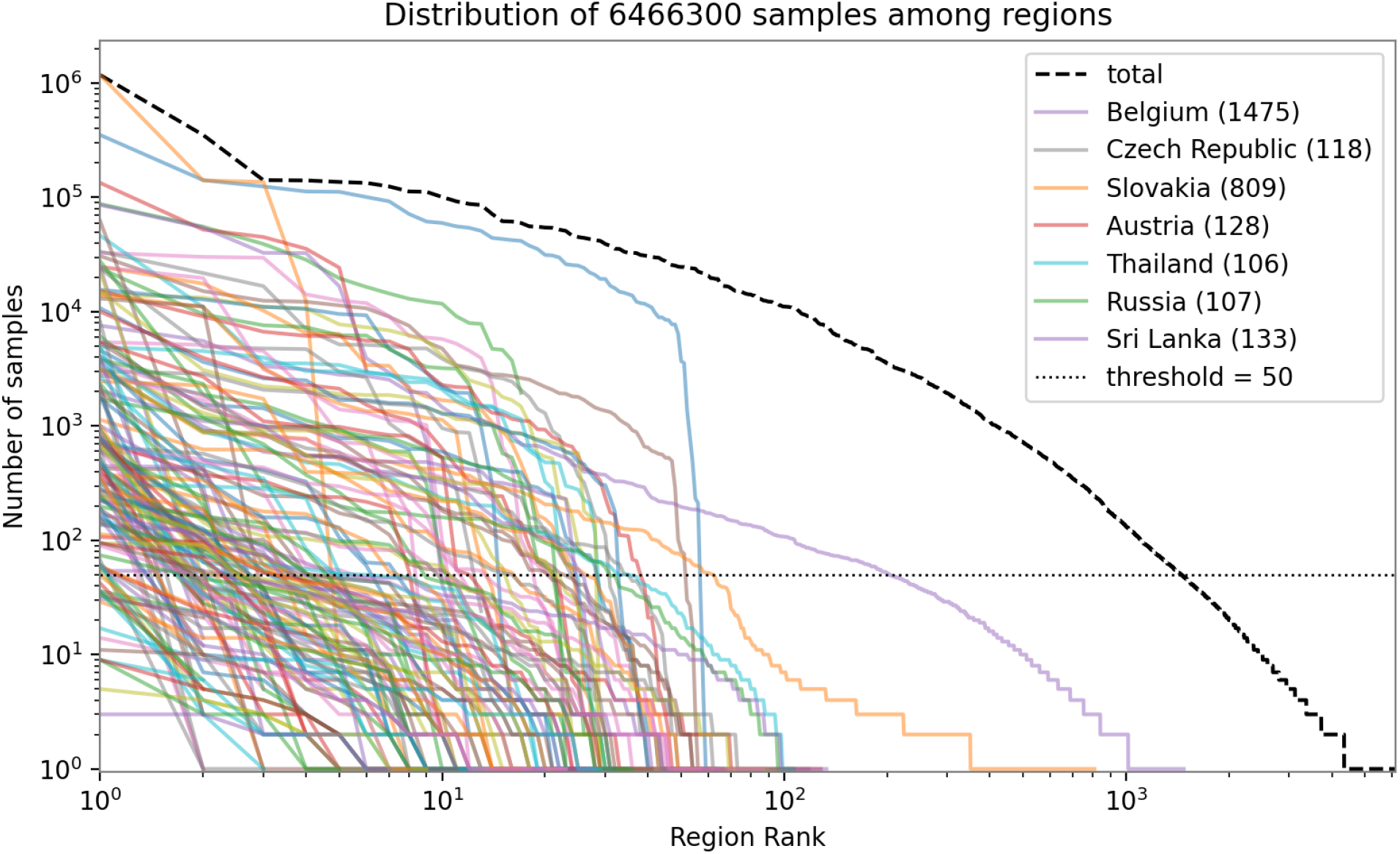
Distribution of samples among regions. Each 2nd level GISAID region (country) is plotted as a curve, with the sizes of 3rd level GISAID regions (usually provinces or states) plotted as points along the curve. The 3rd level is dominated by a few countries with many small regions (e.g. Belgium with 1475 regions), so we merge regions smaller than a threshold (50 samples) into their respective countries.

**Figure S18.**
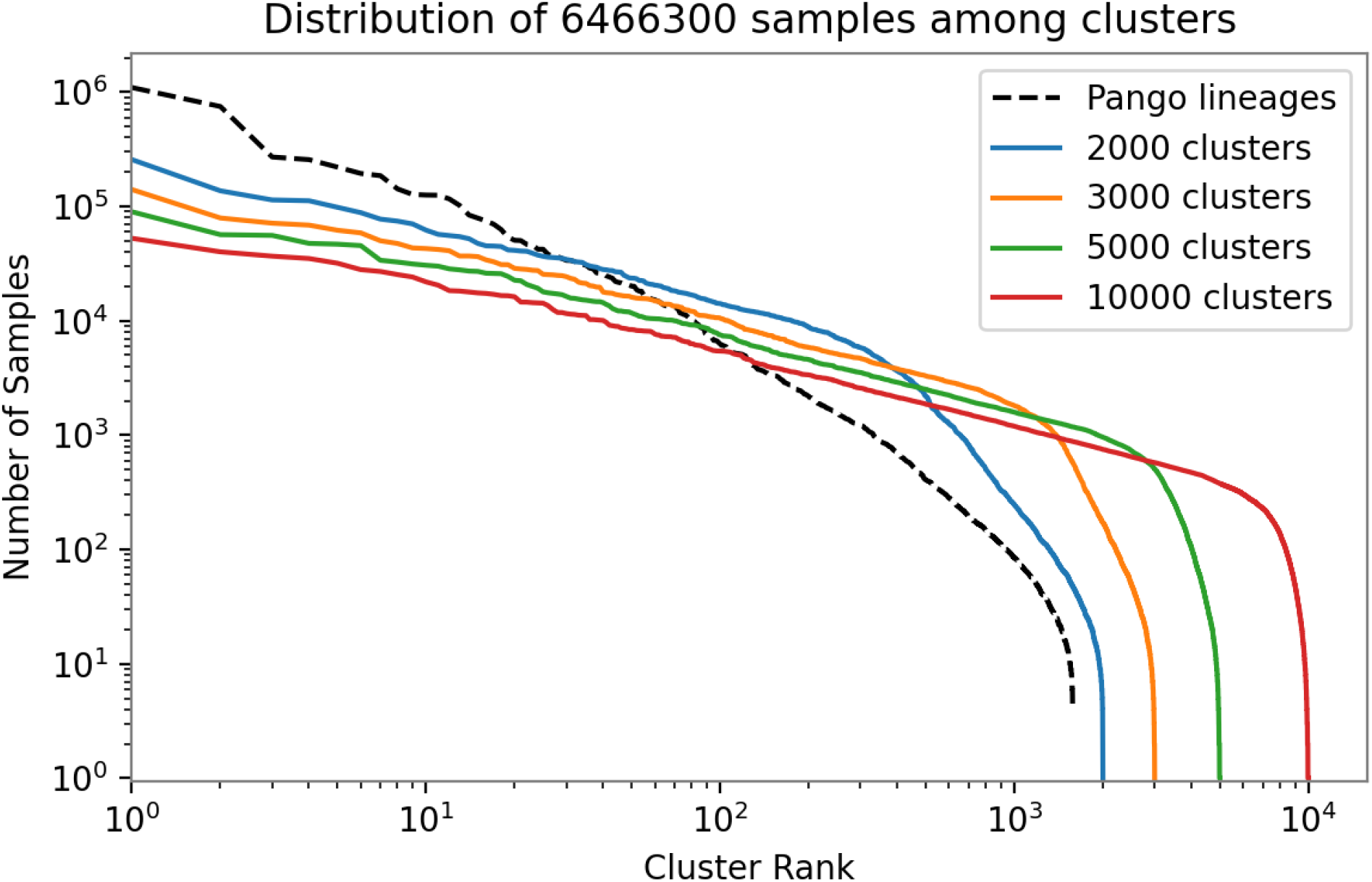
Distribution of samples among PANGO lineages and refined clusters. PANGO lineage sizes are heavy-tailed and appear heterogeneous, so we split into a larger number of clusters (colored). We chose a final clustering of 3000 clusters (orange), balancing between a smaller number of clusters (which improves statistical efficiency) and a larger number of clusters (which better represents lineage heterogeneity).

**Figure S19:**
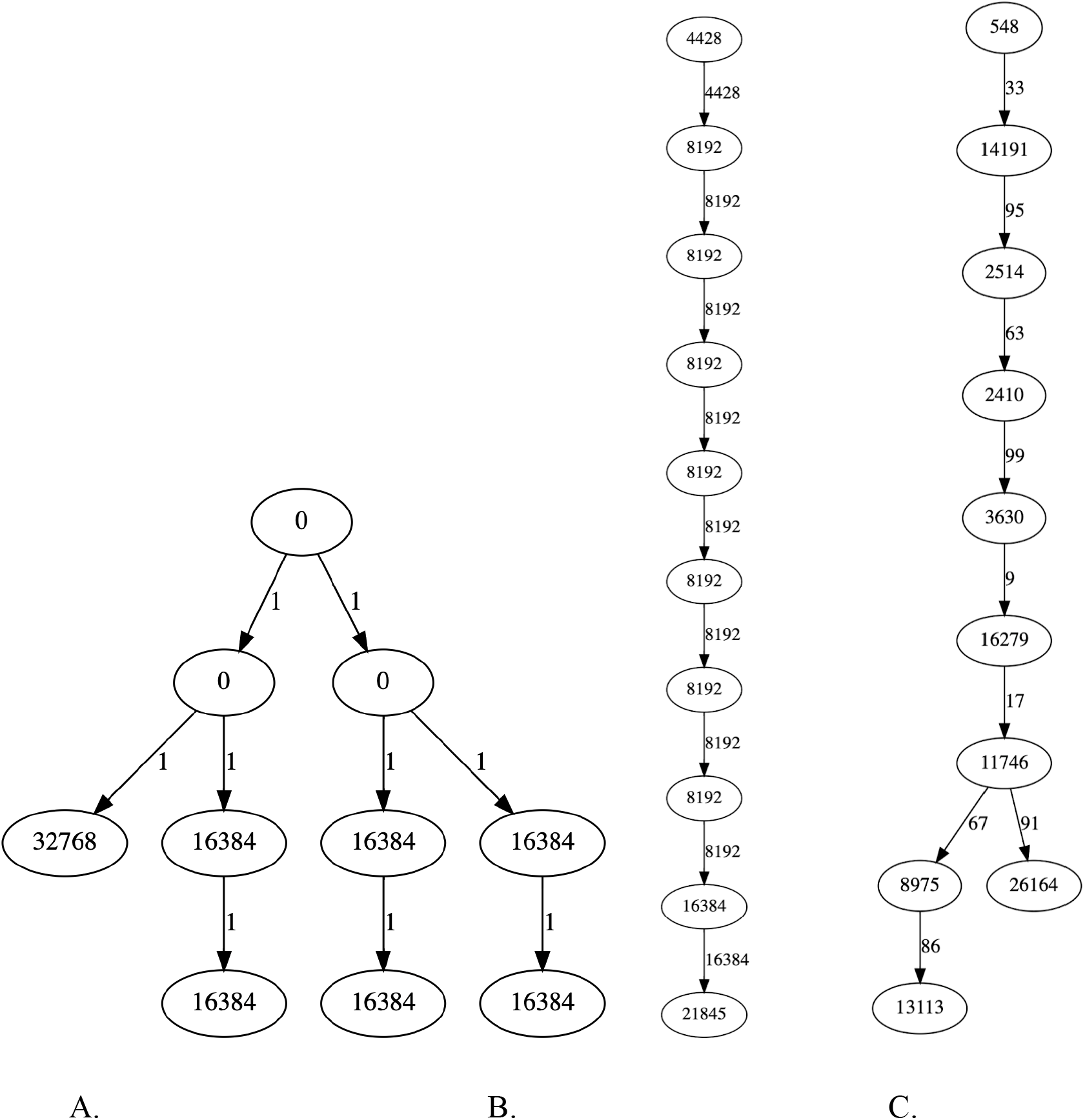
Example algorithmic clustering of three synthetic trees. Example synthetic mutation annotated trees are clustered into smaller trees with only 10 nodes. Nodes are annotated by the number of sequences represented by each cluster. Edges are annotated by the edit distance between clusters. (A) clusters a balanced binary tree of 262,143 nodes, (B) clusters a single linear chain of 20,001 nodes, (C) clusters a random binary tree with Geometric(½)-many children at each node and 200,000 nodes. In all examples the clustered trees are approximately balanced insofar as they exhibit narrow distributions of edge distances (in SNPs) and the cluster sizes (in genomes sampled).

**Figure S20.**
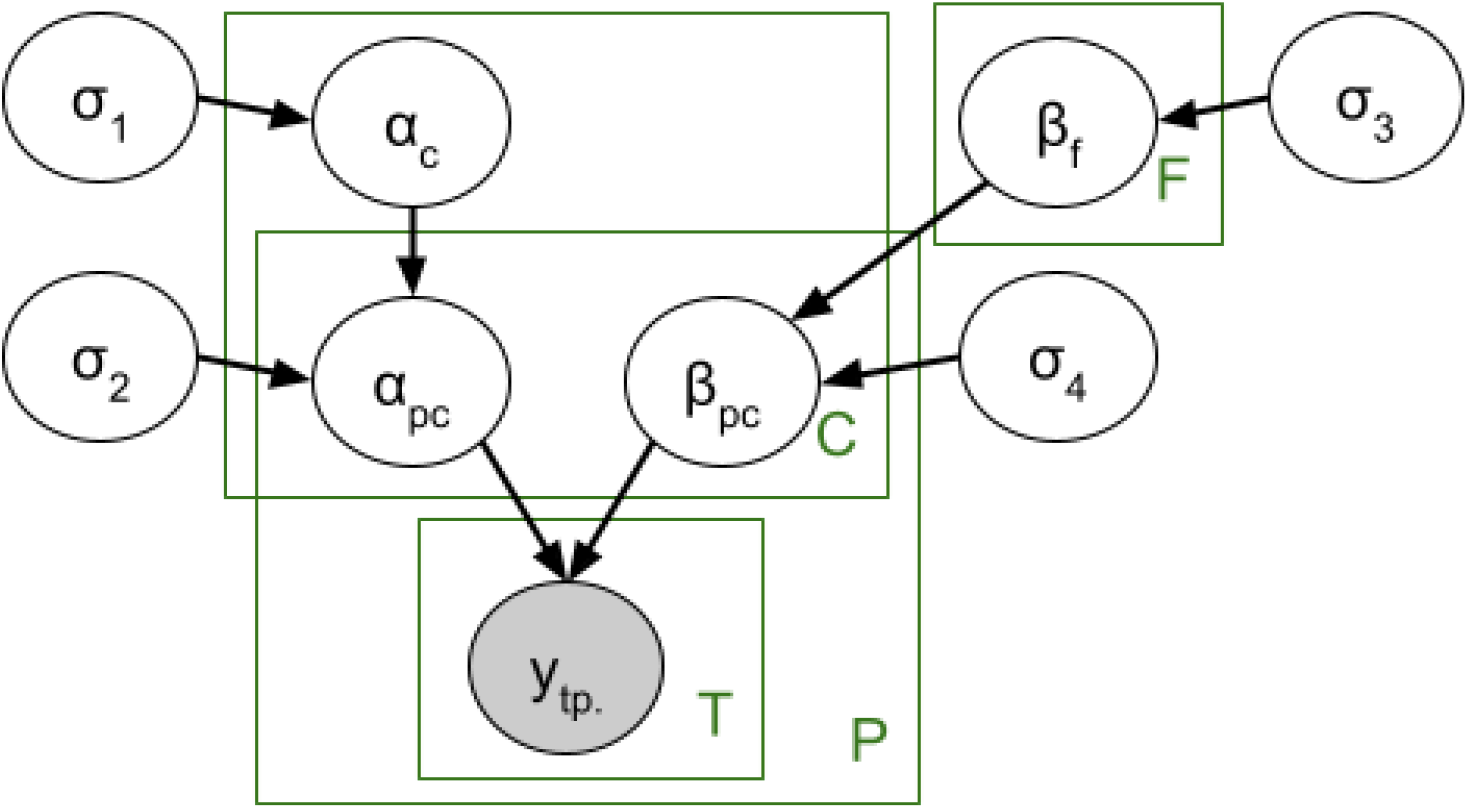
Probabilistic graphical model structure of the PyR_0_ model. Variables σ are scale parameters of distributions. Variable β_f_ is the per-amino-acid-substitution fitness coefficient. Variables β_pc_ and α_pc_ are respectively the per-region per-cluster slope and intercepts parametrizing multivariate logistic growth curves. The mean parameter of β_pc_ is determined by β_f_ via matrix multiplication by the feature matrix X_cf_. The mean parameter of α_pc_ is a per-cluster intercept α_c_ shared across regions. The multinomial observations are vectors y_tp._ each of whose entries y_tpc_ is the number of samples of cluster c in place p in time bucket t. Green boxes denote plates, i.e. conditionally independent replicas of random variables. Note the vector-valued observation y_tp._ is outside of the C plate because the multinomial distribution couples entries across the cluster coordinate c. Because the P x C plates are sparse (in most places most clusters never appear) the model omits α_pc_ and β_pc_ for pairs (p,c) with no observations in y.

**Figure S21.**
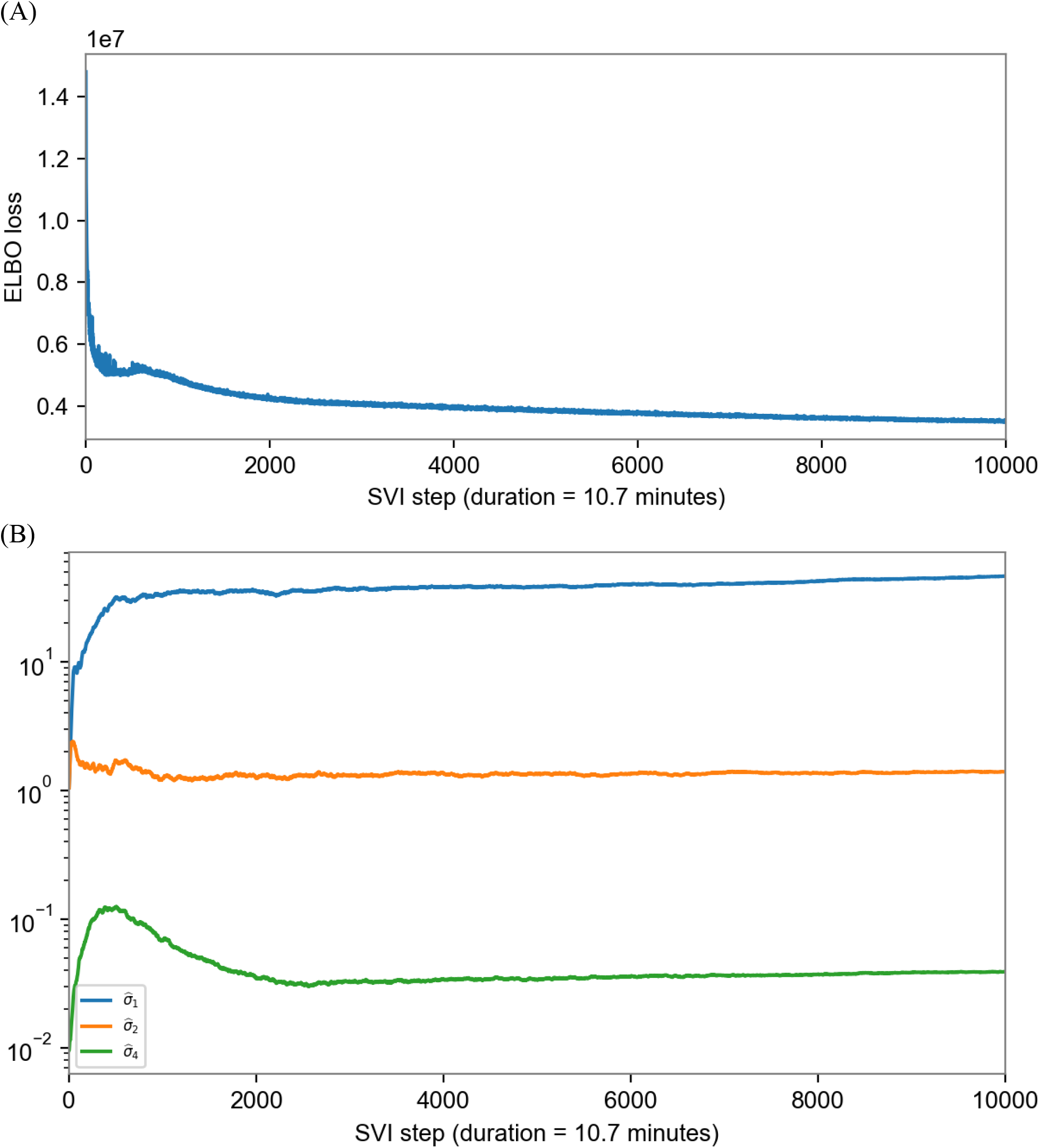
Convergence of variational inference algorithm. **A.** Convergence of ELBO loss. **B.** Convergence of posterior medians of scale parameters.

**Figure S22:**
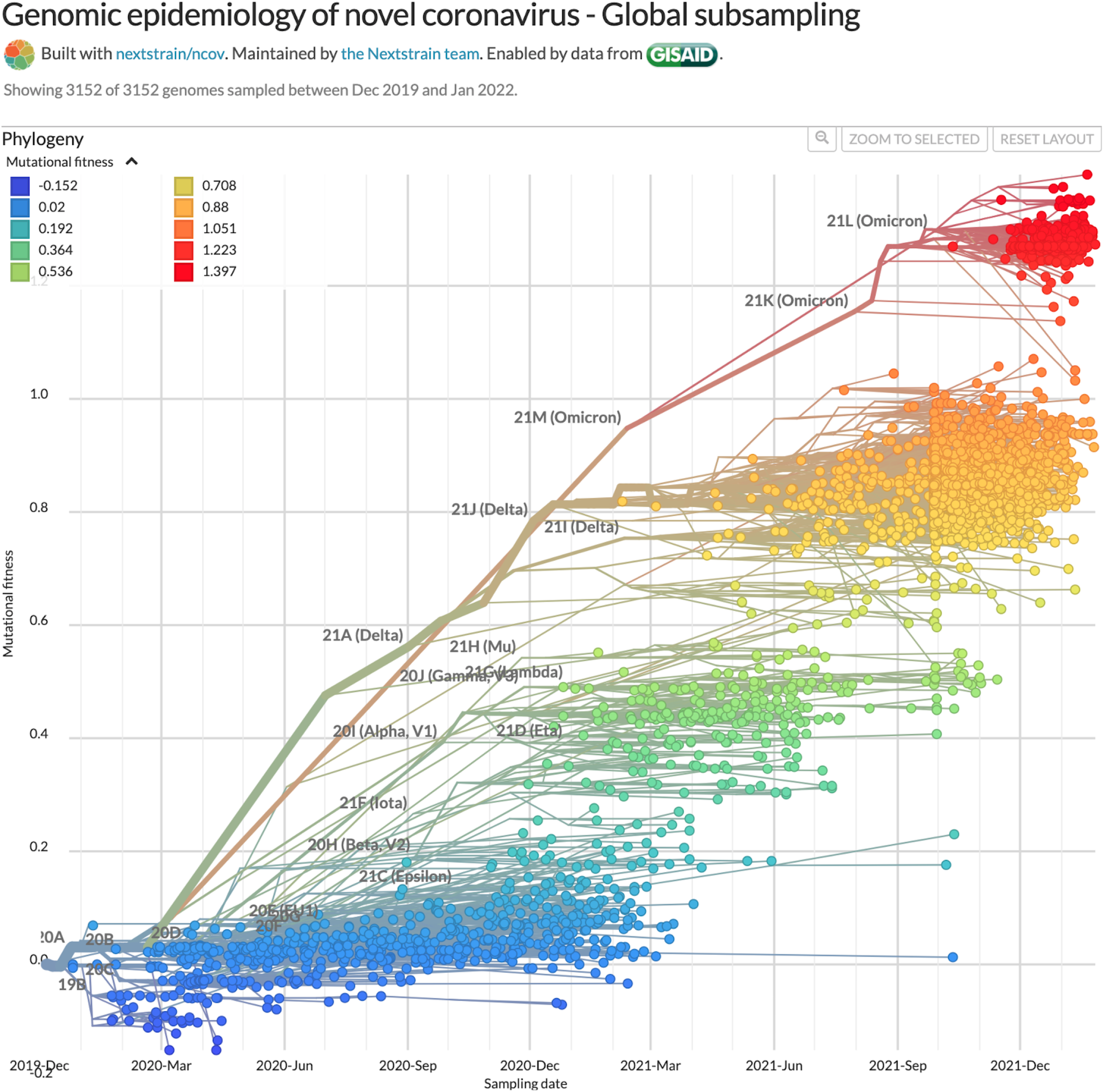
Screenshot of https://nextstrain.org displaying Nextstrain’s subsampled phylogeny with color and y-axis (mutational fitness) determined by our model predicted Δ log R for each lineage (here using a slightly older version of our model). Although PyR_0_ does not explicitly rely on phylogenetic information, fitness estimates vary smoothly across the phylogeny.

**Figure S23.**
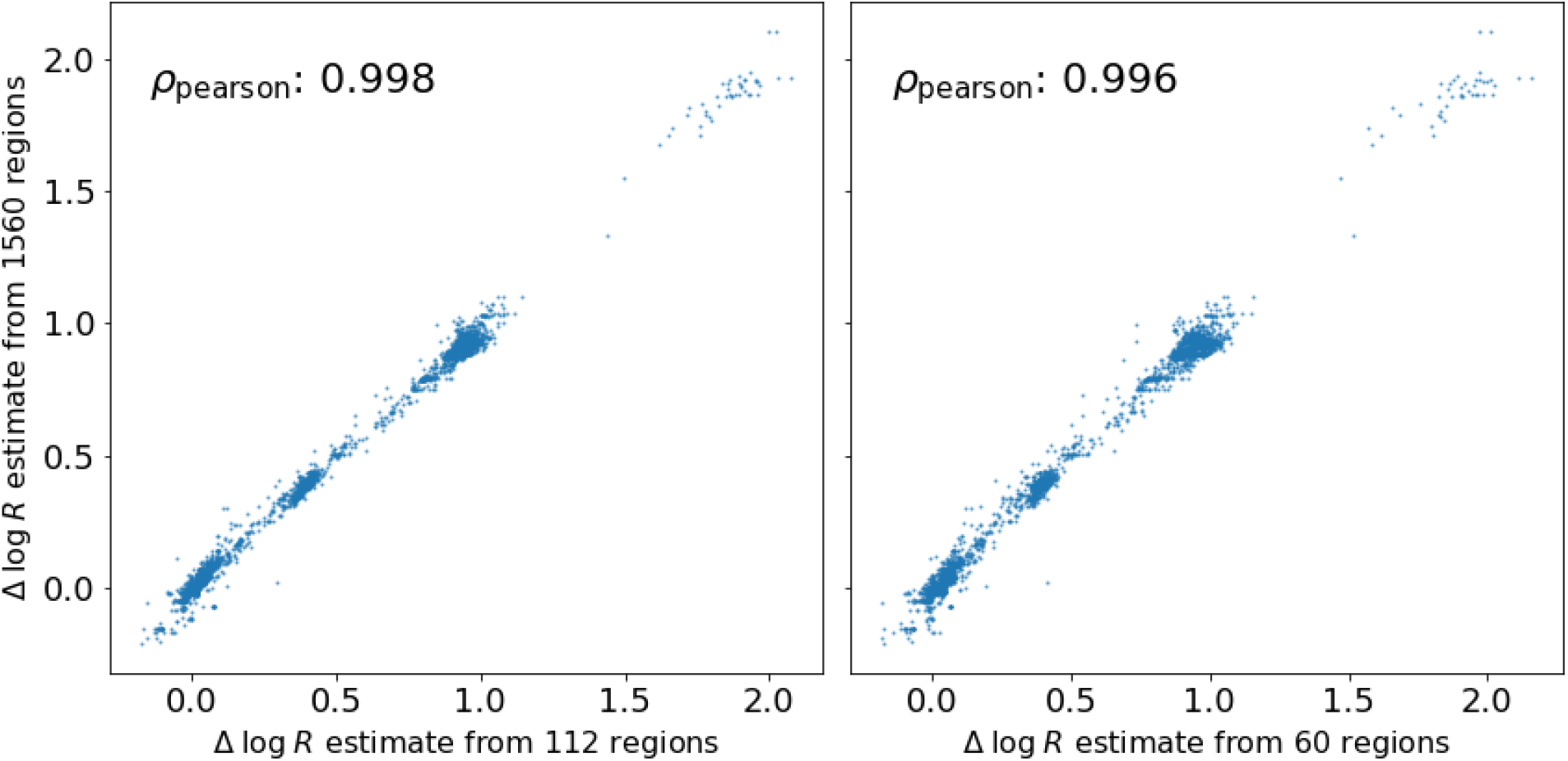
We show that PyR_0_ lineage-level Δ log R estimates are largely driven by regions with the largest numbers of samples, as would be expected from a Bayesian hierarchical model. The vertical axis depicts Δ log R estimates based on all 1560 regions, while the horizontal axis on the left (respectively, right) depicts Δ log R estimates based on the 112 (60) regions with at least 10^4^ (2 ⨉ 10^4^) samples. Collectively these regions contain 80.7% (69.8%) of the total number of SARS-CoV-2 sequences in our full dataset.

**Figure S24.**
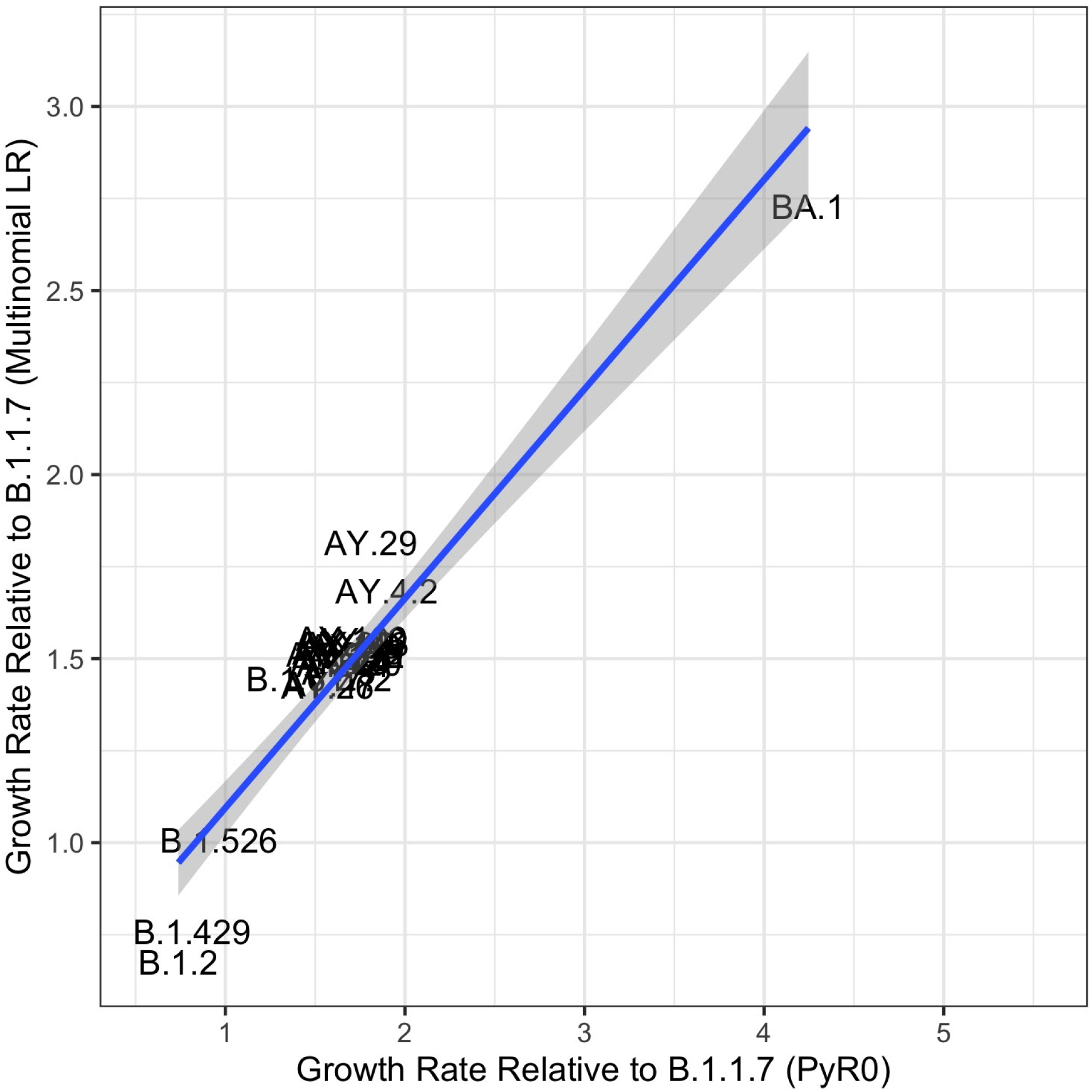
Comparison of mutation-level regression coefficients for growth rate among 50 most prevalent lineages using a standard multinomial logistic regression model with estimates of lineage growth rates from PyR_0_. Pearson’s R = 0.95.

**Figure S25.**
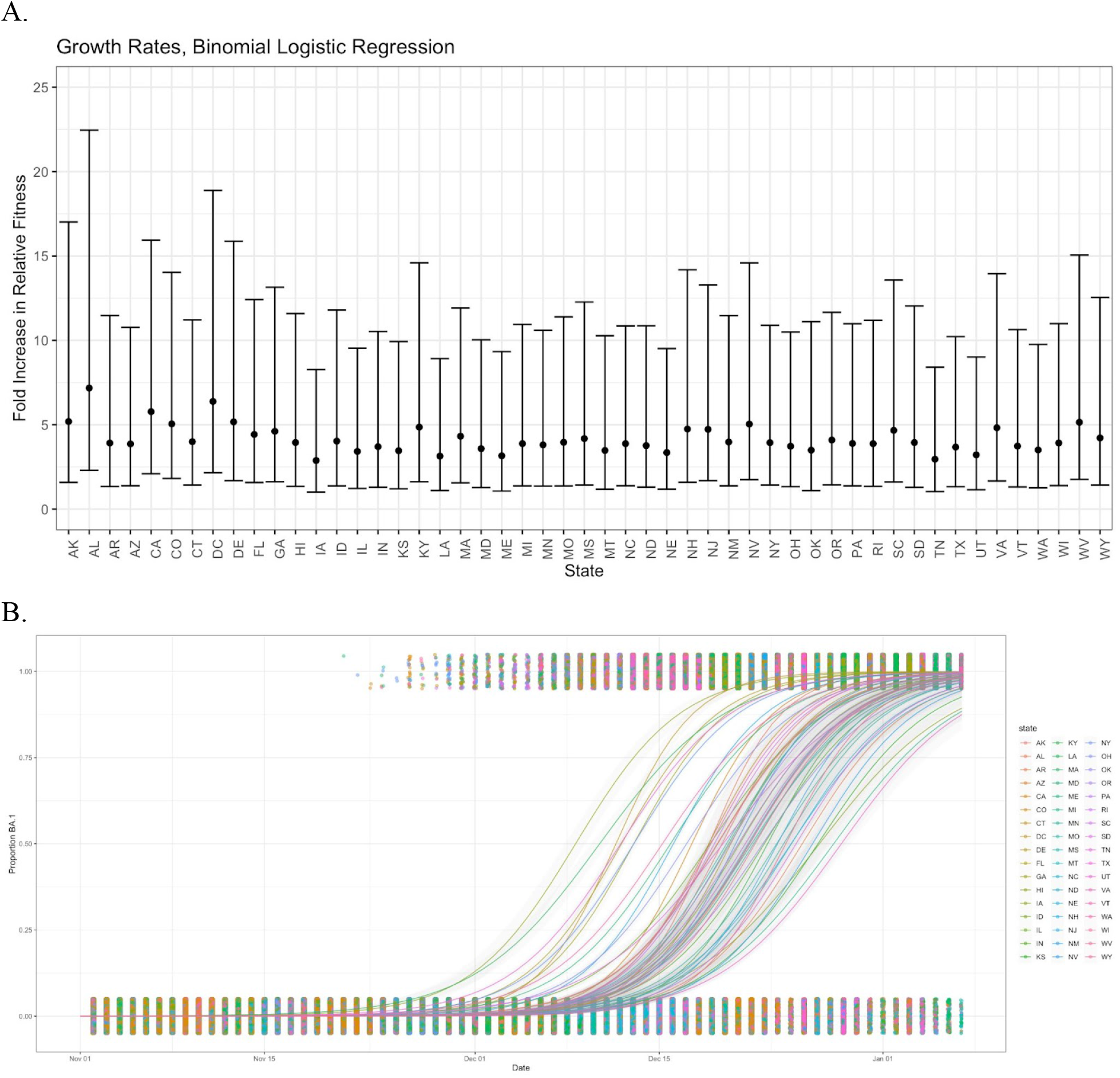
**A.** Estimated growth rate per viral generation (5.5 days) in each state using binomial logistic regressions for the emergence of BA.1 against a background consisting of Delta (B.1.617.2 and sublineages) viruses in all 50 US states between November 1 2021 and January 7 2022. Fold increase in relative fitness is expressed as exp(*β*_1_), where time is measured in viral generations. Error bars show exp(*β*_1_ +/- SE(*β*_1_)).For all 50 US states, the median growth rate per viral generation of Omicron over Delta was 3.9. For all states, the confidence interval for the binomial logistic regression coefficient contained the estimate for the ratio of Omicron to Delta from the PyR_0_ model, which was 3.1 for BA.1.1 / B.1.617.2 and 2.8 for BA.1 / B.1.617.2. **B.** Estimated probability of BA.1 by state from the binomial logistic regression.

**Table S1.** Regional evaluation of forecasts. We evaluate the ability of PyR_0_ to accurately forecast the dominant lineage 4- and 8-weeks into the future in six selected regions with a relatively large number of GISAID samples. Percentage accuracies are obtained by averaging over 45 training windows.

| Region | 4-week forecast | 8-week forecast |
| --- | --- | --- |
| USA | 82.9% | 68.3% |
| France | 75.6% | 61.0% |
| England | 75.6% | 58.5% |
| Brazil | 65.9% | 51.2% |
| Australia | 56.1% | 39.0% |
| Russia | 73.2% | 68.3% |

**Table S2.** Spatial structure of the inferred amino acid coefficients β_f_. We report one-sided p-values for the Moran I spatial autocorrelation statistic computed using a permutation test. We use a gaussian weighting function of the form exp(-distance^2^/lengthscale^2^), where distance is measured in units of nucleotides. We find that there is significant evidence for spatial structure in S, N, ORF7a, ORF3a, and ORF1a as well as across the SARS-CoV-2 genome as a whole.

| Spatial region | # of mutations | Extent of region (nt) | p-value | Lengthscale |
| --- | --- | --- | --- | --- |
| Entire genome | 2904 | 29394 | 0.000001 | 100 |
| Entire genome | 2904 | 29394 | 0.000001 | 500 |
| S | 415 | 3786 | 0.001910 | 50 |
| N | 220 | 1251 | 0.017627 | 50 |
| ORF7a | 75 | 360 | 0.024066 | 18 |
| ORF3a | 198 | 789 | 0.024307 | 39 |
| ORF1a | 1107 | 13182 | 0.029710 | 50 |
| ORF7b | 26 | 126 | 0.089589 | 6 |
| ORF14 | 69 | 213 | 0.112527 | 11 |
| ORF6 | 19 | 177 | 0.138634 | 9 |
| ORF1b | 552 | 8052 | 0.329416 | 50 |
| E | 17 | 195 | 0.455606 | 10 |
| M | 42 | 639 | 0.518497 | 32 |

**Table S3.**
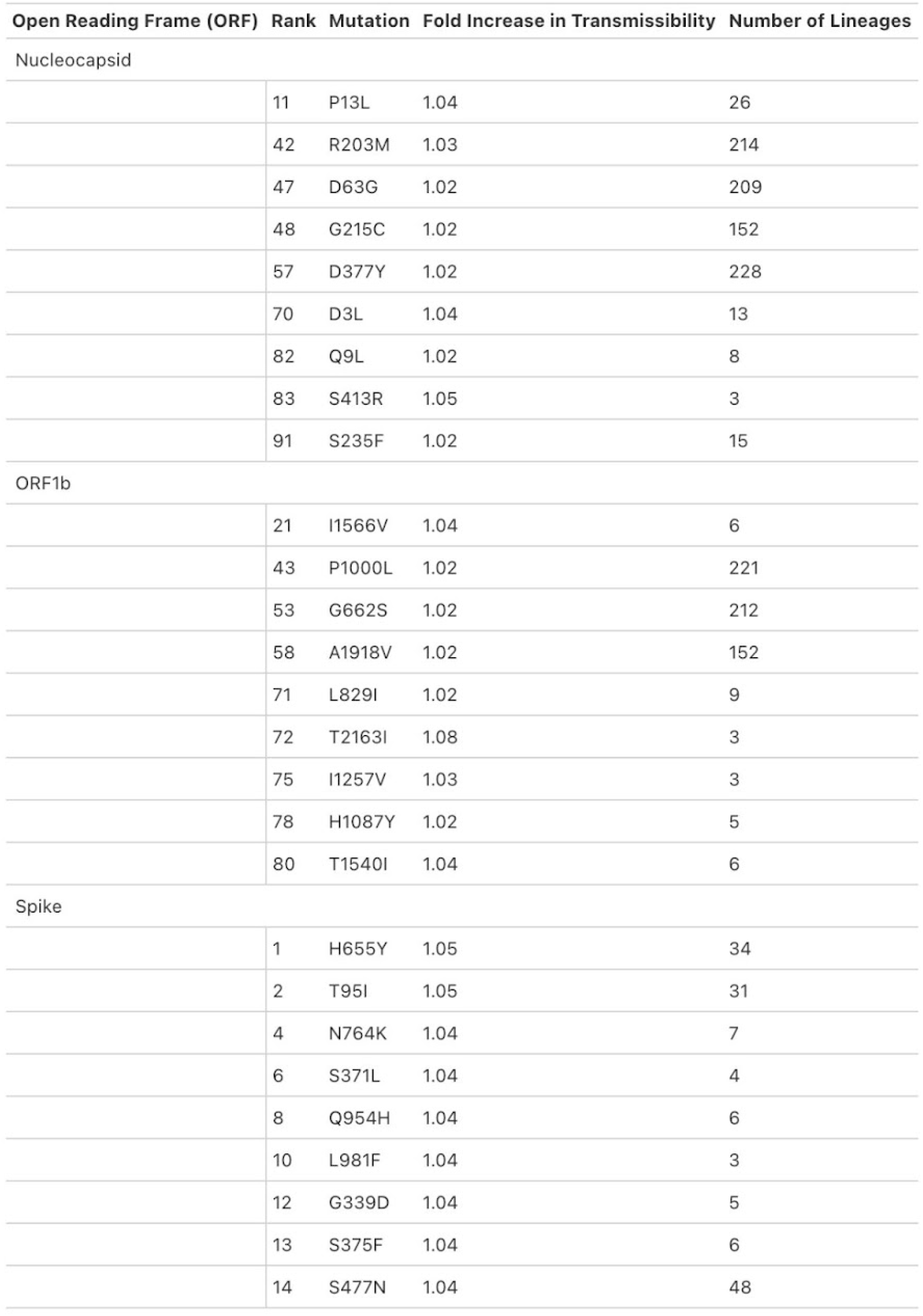
Table of the most statistically significant mutations in spike, ORF1b, and nucleocapsid. The top 9 mutations for each of the listed ORFs is shown. Mutations such as S:H655, S:T95I, and N:R203M have emerged independently in VoC lineages.

**Data S1.** (separate file strains.tsv)

Complete list of PANGO lineages with inferred relative fitness, ranked by fitness. Mirrored at https://github.com/broadinstitute/pyro-cov/blob/v0.2/paper/strains.tsv

**Data S2.** (separate file mutations.tsv)

Complete list of amino acid changes with inferred effect size, ranked by z-score. Mirrored at https://github.com/broadinstitute/pyro-cov/blob/v0.2/paper/mutations.tsv

**Data S3.** (separate file accession_ids.txt.xz)

Complete list of GISAID accession numbers of viral genomes used in this study.

Mirrored at https://github.com/broadinstitute/pyro-cov/blob/v0.2/paper/accession_ids.txt.xz

## Acknowledgements

We acknowledge crucial assistance in data preprocessing from Angie Hinrichs. We thank Trevor Bedford and Cornelius Roemer for visualizing the outputs of our model on nextstrain.org. We acknowledge helpful discussions and feedback from Du Phan, William Hanage, Christopher Tomkins-Tinch, Shira Weingarten-Gabbay, Katie Siddle, Sagar Gosai, Steven Reilly, Eli Bingham, Mehrtash Babadi, Holly Soutter, Debora Marks, Noor Youssef, Sarah Gurev, and Nicole Thadani. We gratefully acknowledge the authors from the originating laboratories and the submitting laboratories, who generated and shared via GISAID genetic sequence data on which this research is based.

## Author contributions

Conceptualization: F.O., S.F.S., J.E.L., M.J.

Data curation: F.O., N.B.

Formal Analysis: F.O., S.F.S, M.J., N.B., J.E.L.

Funding acquisition: B.M., P.C.S, J.L., J.E.L.

Investigation: all authors

Methodology: F.O., S.F.S, M.J., J.E.L., L.Y., M.B.

Project administration: all authors Software: F.O., N.B., M.J.

Supervision: D.J.P., B.M., J.L., P.C.S., J.E.L.

Validation: F.O., N.B., M.J., S.F.S.

Visualization: F.O., J.E.L., N.B., J.P., S.F.S.

Writing – original draft: F.O., S.F.S., B.M., P.C.S, J.E.L.

Writing – review & editing: all authors

Authors have no competing interests.

## Data and materials availability

We gratefully acknowledge all data contributors, i.e. the Authors and their Originating laboratories responsible for obtaining the specimens, and their Submitting laboratories for generating the genetic sequence and metadata and sharing via the GISAID initiative (*13*) on which this research is based. A total of 6,466,300 submissions are included in this study. A complete list of 6.4million accession numbers is included as Data (S3).

## List of Supplementary materials

Materials and Methods

Supplemental Note 1: Detailed description of PyR0 model

Supplemental Note 2

Supplemental Note 3

Fig S1 – S25

Table S1 – S3

References (20–47)

Data S1 (strains.tsv), S2 (mutations.tsv), S3 (accession_ids.txt.xz)

